# Genome-wide association study identifies *RNF123* locus as associated with chronic widespread musculoskeletal pain

**DOI:** 10.1101/2020.11.30.20241000

**Authors:** Md Shafiqur Rahman, Bendik S Winsvold, S.O. Chavez Chavez, Sigrid Børte, Yakov A. Tsepilov, Sodbo Zh. Shapov, HUNT All-In Pain, Yurii Aulchenko, Knut Hagen, Egil A. Fors, Kristian Hveem, John-Anker Zwart, J.B.J. van Meurs, Maxim B. Freidin, Frances M.K. Williams

**Author notes:** Corresponding Author Frances MK Williams PhD FRCP(E), Professor of Genomic Epidemiology & Hon Consultant Rheumatologist, Department of Twin Research and Genetic Epidemiology, King’s College London, Westminster Bridge Road, London SE1 7EH.

## Abstract

**Background and Objectives:** Chronic widespread musculoskeletal pain (CWP) is a symptom of fibromyalgia and a complex trait with poorly understood pathogenesis. CWP is heritable (48-54%), but its genetic architecture is unknown and candidate gene studies have produced inconsistent results. We conducted a genome-wide association study to get insight into the genetic background of CWP.

**Methods:** Northern Europeans from UK Biobank comprising 6,914 cases reporting pain all over the body lasting more than 3 months and 242,929 controls were studied. Replication of three lead genome-wide significant single nucleotide polymorphisms (SNPs) was attempted in 6 independent European cohorts (N=43,080; cases=14,177). Genetic correlations with risk factors, tissue specificity, and colocalization were examined.

**Results:** Three genome-wide significant loci were identified (*rs1491985, rs10490825, rs165599*) residing within the genes *RNF123, ATP2C1*, and *COMT*. The *RNF123* locus was replicated (meta-analysis p=0.0002), the *ATP2C1* locus showed suggestive association (p=0.0227), and the *COMT* locus was not replicated. Partial genetic correlation between CWP and depressive symptoms, body mass index, age of first birth, and years of schooling were identified. Tissue specificity and colocalization analysis highlight the relevance of skeletal muscle in CWP.

**Conclusions:** We report a novel association of *RNF123* locus with CWP and suggest a role of *ATP2C1*, consistent with a role of calcium regulation in CWP. The association to *COMT*, one of the most studied genes in chronic pain field, was not confirmed in the replication analysis.

**Key messages:** *What is already known about this subject?:*

- Chronic widespread musculoskeletal pain (CWP) is a primary diagnostic feature of fibromyalgia.
- CWP is moderately heritable, but precise genes involved in the pathogenesis of CWP are yet to be identified.

*What does this study add?:*

- This is the largest genetic study conducted on CWP to date and identified novel genetic risk loci (*RNF123* and *ATP2C1*).
- The genetic signal points to peripheral pain mechanisms in CWP, and shows genetic correlation with other traits, including BMI and depression.

*How might this impact on clinical practice or future developments?:*

- The findings add to etiological basis of CWP.

## Introduction

Chronic widespread musculoskeletal pain (CWP) is a common complex trait influenced by genetic and environmental factors, most of which have yet to be determined[1]. CWP and fibromyalgia syndrome are sometimes used interchangeably although the latter is generally more severe and includes other features such as sleep disturbance, fatigue and depression [2]. It is thought to represent a subgroup at the more severe end of the spectrum of CWP[3]. The prevalence of CWP is 10.6% in the world population and 14.2% in the UK population[4, 5]. It is associated with high societal cost[6]. CWP is responsible for excess mortality[7], which is thought to be attributable to cardiovascular disease, respiratory disease and cancer. Females are more affected by CWP than males[4], and the prevalence rises with age[5]. In addition to age and sex, a number of exposures have been proposed as risk factors for CWP[8, 9] but only increased body mass index (BMI) has been consistently reported across studies, including longitudinal studies[10-12].

Heritability estimates for CWP range between 48-54%, indicating a substantial genetic contribution[13]. To date, the candidate gene approach has been extensively applied to identify genetic factors in CWP[14] (see below) but few agnostic studies have been published[15]. The only genome-wide association study (GWAS) meta-analysis combining 14 studies identified a locus lying on chromosome 5 intergenic to *CCT5* and *FAM173B*[15]. *CCT5* has previously been implicated in neuropathy[16] and there is increasing evidence that small fiber neuropathy underlies a subset of fibromyalgia[17].

Genetic factors are known to be shared by chronic pain conditions[18, 19]. One of the most extensively studied chronic pain-associated genes encodes catechol-O-methyltransferase (COMT), an enzyme which regulates the production of catecholamines that act as neurotransmitters in the central nervous system pain tract. A non-synonymous change of A to G encoding a valine (Val) to methionine (Met) substitution at codon 158 (Val158Met; *rs4680*) reduces the enzymatic activity of *COMT*. This single nucleotide polymorphism (SNP) has been reported to be associated with CWP in a small study of 122 participants[20], but a subsequent association study of 3,017 participants did not confirm earlier findings[21]. An inconclusive role of *COMT* was observed for temporomandibular disorders (TMD) as well[22, 23]. Further investigation is required to identify genetic variants underlying CWP which will shed light on the pathophysiological mechanisms underlying the development of chronic pain and may reveal therapeutic targets.

## Materials and Methods

An overview of study design presented in figure 1.

**Figure 1.**
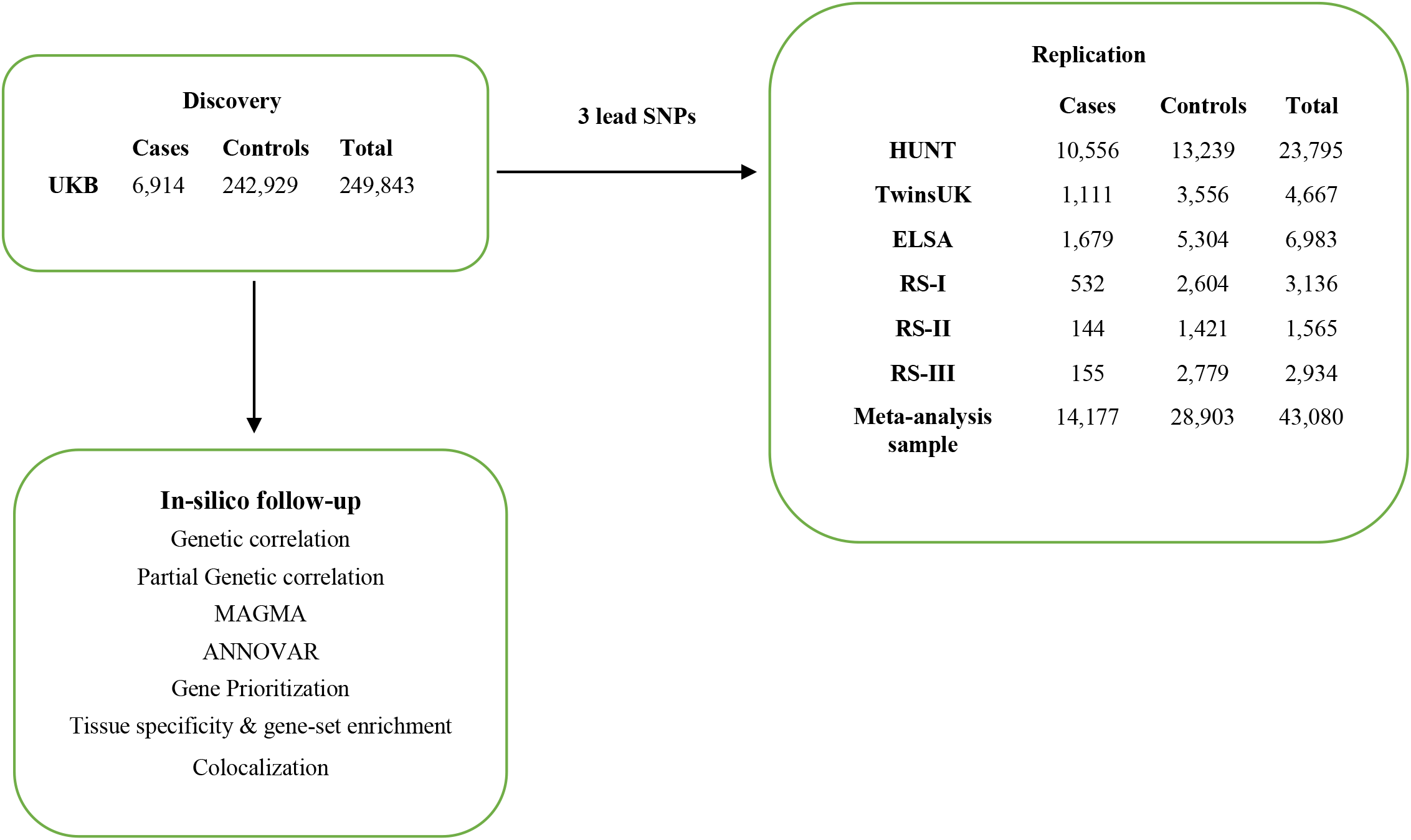
Overview of study design.

### Participant selection

For the discovery analysis, we performed a GWAS of CWP using UK Biobank (UKB) comprising 249,843 participants of European descent (6,914 CWP cases and 242,929 controls). Independent SNPs passing a threshold p<5.0E-08 were submitted for replication in 43,080 individuals of European ancestry (14,177 CWP cases and 28,903 controls) from six independent cohorts originating in the UK (TwinsUK and The English Longitudinal Study of Ageing (ELSA)), the Netherlands (The Rotterdam Study I, II and III (RS-I, RS-II and RS-III)), and Norway (The Nord-Trøndelag Health Survey (HUNT)). All study participants provided written informed consent. Study-specific ethical approval was provided by the local ethics committees. The UKB dataset was used under project #18219. Description of each study cohort is presented in supplementary methods.

### Phenotype

In UKB, CWP cases were defined by combining self-reported diagnosis of pain all over the body lasting for more than 3 months; simultaneous pain in the knee, shoulder, hip and back lasting 3+ months; and doctor’s diagnosis of fibromyalgia. Controls comprised those who reported no pain in the last month or reported pain all over the body in the previous month that did not last for three months or reported only ≥ 3 months of non-musculoskeletal pain (headache, facial and abdominal pain). Those reporting a diagnosis of rheumatoid arthritis, polymyalgia rheumatica, arthritis not otherwise specified, systemic lupus erythematosus, ankylosing spondylitis or myopathy were excluded from the study (supplementary figure S1). Further phenotype details for UKB and replication cohorts are provided in supplementary methods.

### Genotyping

Genotyping and imputation methods across cohorts are summarised in supplementary table S1. In brief, UKB participants were genotyped using Applied Biosystems UKB Axiom array and Applied Biosystems UKB Lung Exome Variant Evaluation Axiom array. HUNT participants were genotyped using Illumina/HumanCoreExome12 v1.0, Illumina/HumanCoreExome12 v1.1 and UM HUNT Biobank v1.0. ELSA participants were genotyped using the Illumina/HumanOmni2.5-4v1 and Illumina/HumanOmni2.5-8v1.3. TwinsUK participants were genotyped using Illumina/HumanHap300, Illumina/HumanHap610Q, Illumina/1M-Duo and 1.2MDuo 1M. RS-I participants were genotyped using Illumina/HumanHap 550K V.3 and Illumina/HumanHap 550K V.3 DUO. RS-II participants were genotyped using Illumina/HumanHap 550V.3DUO and Illumina/HumanHap610Q. RS-III genotyping was performed using Illumina/HumanHap610Q.

### Statistical analysis and in-silico follow-up

The details of statistical analysis, and in-silico follow-up are described in supplementary methods. In brief, GWAS in the discovery sample was performed using linear mixed-effects model implemented in BOLT-LMM v2.3.2[24]. An additive genetic model for SNP effect on CWP was adjusted for age, genetically determined sex, genotyping platform, and the first ten genetic principle components[25]. A sensitivity GWAS (controls: 223,606 and CWP cases: 6,914) was performed excluding participants with chronic non-musculoskeletal pain such as headache, facial and abdominal pain from the controls. Independent SNPs at GWAS significant loci were identified using Conditional & Joint[26] (COJO) analysis and submitted for replication. Independent SNPs across all replication cohorts were meta-analysed using fixed-effects model with both sample size, and inverse-variance weighting implemented in METAL[27]. LD score regression (LDSR)[28] was used to estimate inflation (λGC) in test statistics, SNP-based heritability on the observed and liability scale and genetic correlations. We also estimated partial genetic correlations[29]. We used FUMA[30] for the functional annotation of SNPs, gene mapping, tissue specificity and gene-set enrichment. Differential expression of replicated lead SNP was assessed using the GTEx version-8 tissues. Colocalization of GWAS independent SNPs in human skeletal muscle and dorsal root ganglion (DRG) tissues was assessed utilizing publicly available data[31, 32].

## Results

Details of the discovery and replication cohorts are presented in table 1. Cases were enriched for females compared to controls in all cohorts (p<0.001) and were on average older in the discovery, and in four replication cohorts (p<0.05). In all cohorts BMI was significantly higher in cases than controls (p<0.0001) except for RS-3 where a similar but non-significant trend was observed (p=0.0827).

**Table 1.**
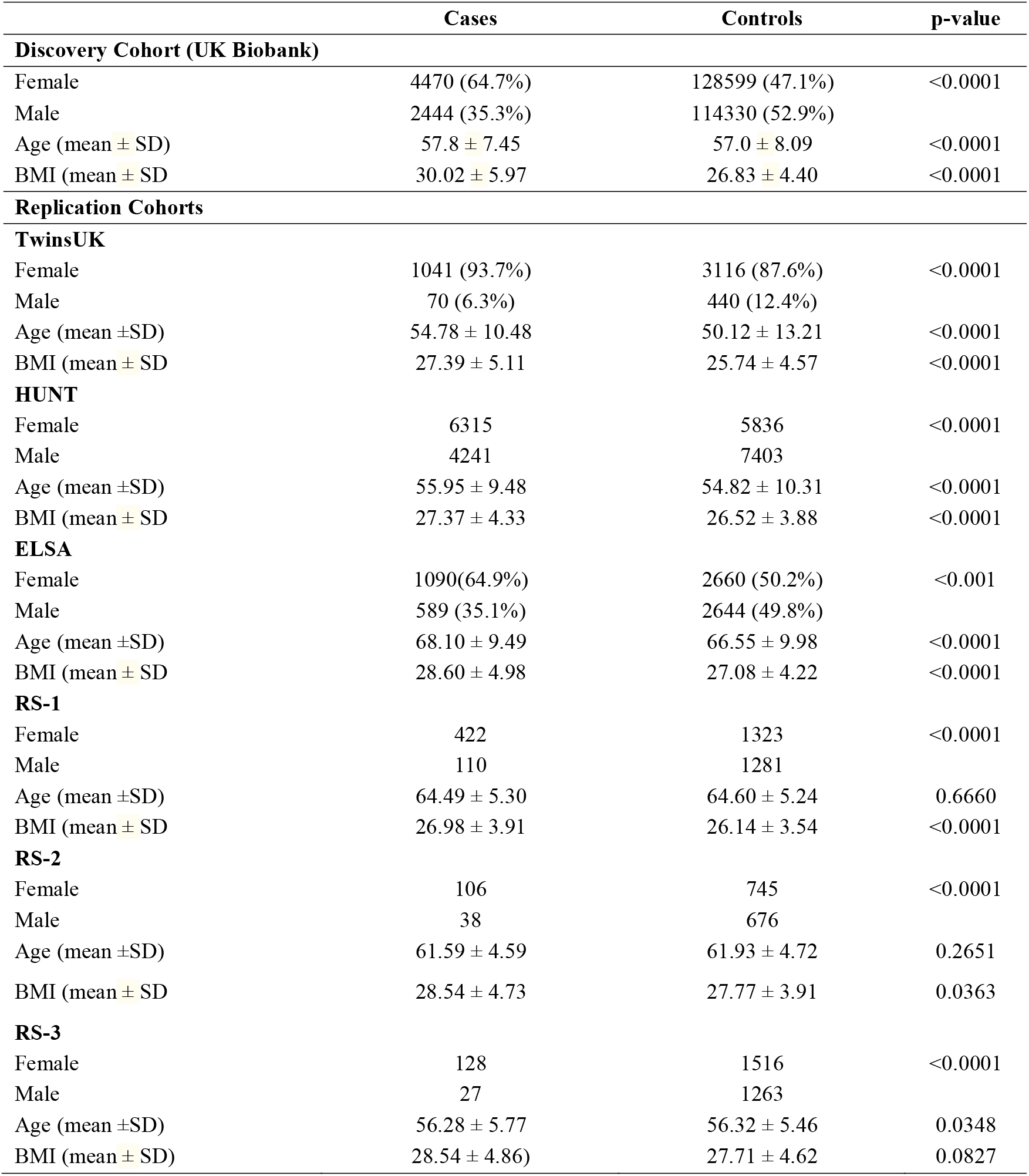
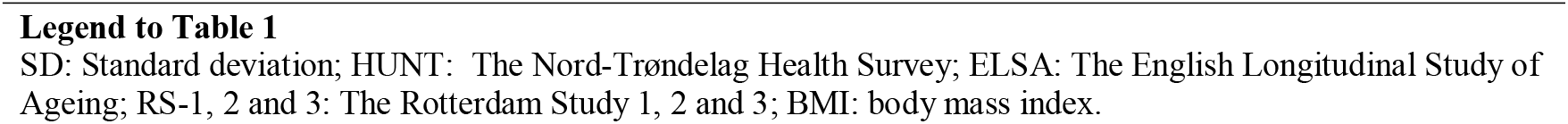
Sample characteristics stratified by case / control status for discovery and replication cohorts.

### Discovery genome-wide association study

Three genomic loci tagged by *rs1491985, rs10490825*, and *rs165599* passed genome-wide significance threshold of p<5E-08 (figure 2). Observed inflation in test statistics (λGC = 1.146, supplementary figure S2) was due to polygenicity (LDSR intercept: 1.002±0.0085, LDSR ratio=0.0118±0.0497) rather than confounding. SNP-heritability of CWP was 0.033±0.004 on the observed scale, and 0.22±0.02 on the liability scale meaning that the observed SNPs explain 22% of variance in CWP risk. Lead SNPs were located in the gene *RNF123* (chromosome 3, *rs1491985*, intronic variant, p=1.60E-08), *ATP2C1* (chromosome 3, *rs10490825*, intronic variant, p=1.30E-08), and *COMT* (chromosome 22, rs165599, 3’-UTR variant, p=2.50E-08), respectively (figure 3A,C; supplementary table S2). Six additional loci near or within genes *HNRNPA1P46, LRRC3B, PDE6A, DPYSL2, ANXA11*, and *AL138498*.1 were identified at suggestive GWAS threshold of p<5E-07. Sensitivity GWAS excluding participants with chronic non-musculoskeletal pain provided findings similar to original results except that *COMT* locus now became suggestively significant (p=5.3E-08) (supplementary figure S3).

**Figure 2.**
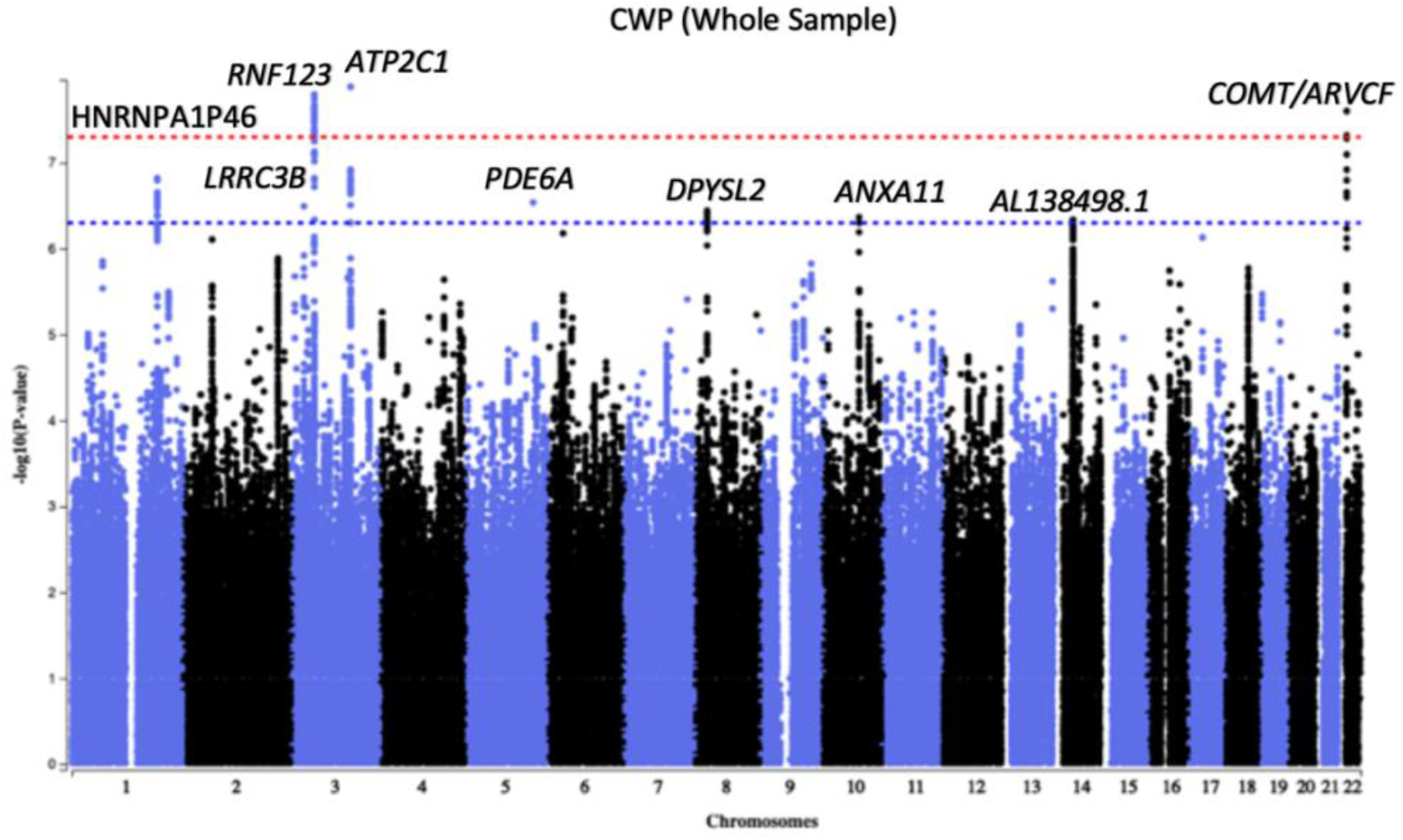
Manhattan plot of summary GWAS of chronic wide-spread pain (CWP) derived from UK biobank European ancestry data. Each circle in the plots represents an SNP, which was positioned following genomic build GRCh37. Colored circles indicated linkage disequilibrium (LD). The y-axis shows the corresponding –log10 P-values based on UKB discovery cohort and the x-axis show chromosome positions along with gene names and flaking size.

**Figure 3.**
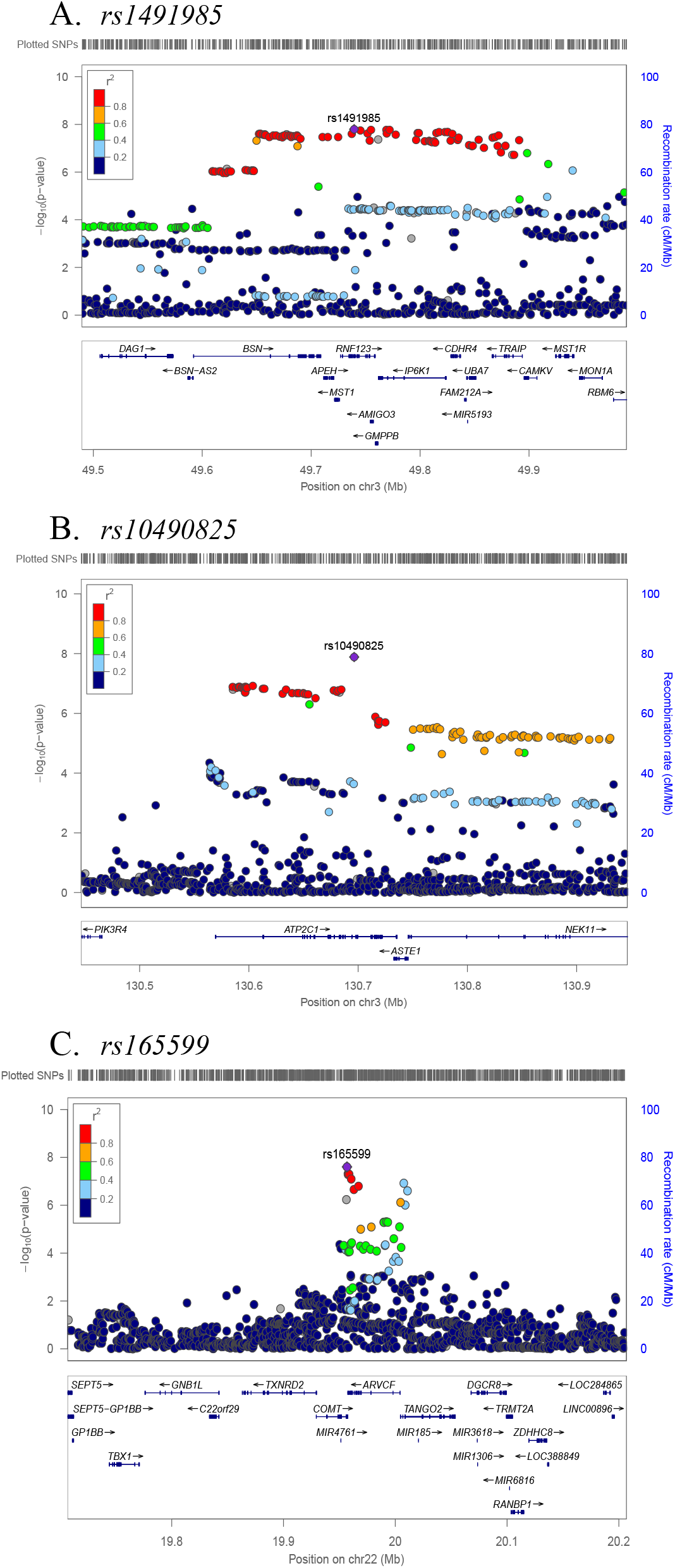
Regional plots for three independent CWP associated SNPs. Independent SNPs are colored in purple. Other colored circles indicated pairwise LD. Strength of LD (r^2^) presented in the upper right corner of each plot.

### Replication results and meta-analysis

Results are presented in supplementary table S3, with meta-analysis of the 6 replication samples in figure 4 (supplementary table S4,S5). Given the significance threshold for replication: 0.05/3=0.017, association between CWP and *rs1491985* was considered replicated (sample-size based p=0.0002; standard-error based p=0.0003). *Rs10490825* showed suggestive association with CWP (sample-size based p=0.0227; standard-error based p=0.0490) and demonstrated a consistent direction of effect in 5 of the 6 replication samples. *Rs165599* did not replicate (sample-size based p-value=0.7300; standard-error based p-value=0.5000) and the direction of effect was not consistent across cohorts: in three cohorts, allele A was protective, while in the other three it was the risk allele. None of the three SNPs displayed statistically significant heterogeneity in the replication cohorts.

**Figure 4.**
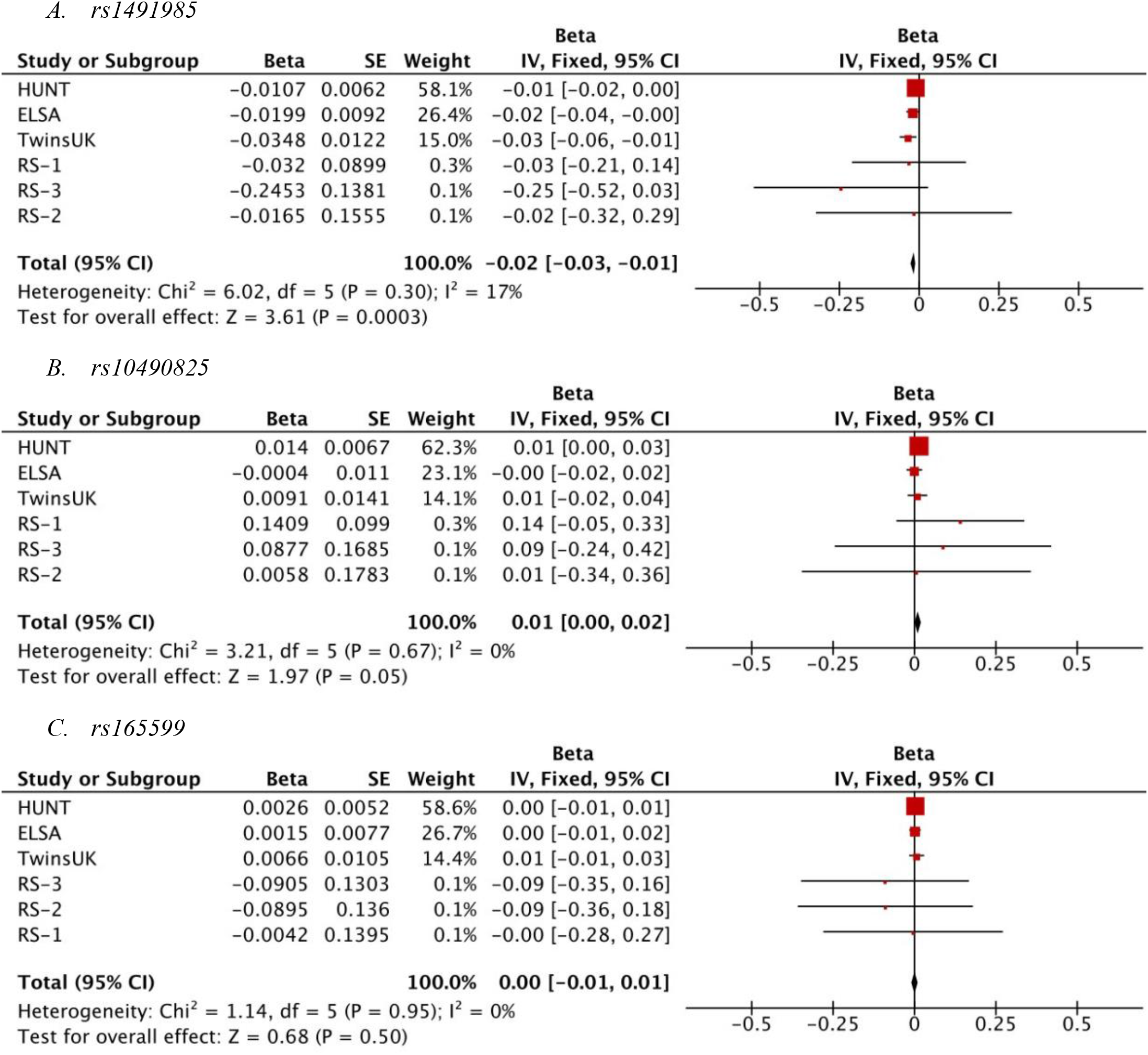
Forest plot of the association for (A) *rs1491985*, (B) *rs10490825*, and (C) *rs165599* with chronic widespread pain (CWP). X-axis shows, effect size measures are presented as beta value. The red square with horizontal black line represents the cohort-specific effect with a corresponding confidence interval for the SNP of interest. Size of the square indicates the weight of the study and reflects sample size. The vertical black line indicates “line of no effect”. Overall effect is presented as a black diamond. Test-statistics for each cohort, meta-analysis and heterogeneity are available on the left-hand side. The *rs1491985*, and *rs10490825* were not present in ELSA, therefore the *rs9870858*, and *rs1732984*8 were used as proxy, respectively (online supplementary methods).

### CWP shares genetic components with BMI, depression, age at first birth, and years of schooling

Two hundred and nine traits from sources other than UKB were examined for genetic correlation with CWP. We selected traits for which the absolute value of the correlation coefficient (rg) was >0.2, and for which the Bonferroni-corrected p-value was <0.01/209=4.78E-05. Twenty-three traits fulfilled these criteria (supplementary figure S4). The highest positive genetic correlation was observed for depressive symptoms (rg=0.65) and the highest negative correlation was observed for college completion (rg= −0.61). Many of the 23 genetically correlated traits were correlated with each other raising concerns about their independency of correlations with CWP. We therefore calculated partial genetic correlations conditionally independent of each other.

Using hierarchical clustering, based on genetic correlations we identified 7 clusters (supplementary figure S5A), with 7 traits selected to represent each cluster (BMI, triglycerides, depressive symptoms, coronary artery disease, smoking, age of first birth, and years of schooling) to quantify partial genetic correlation with CWP. We found depressive symptoms (rg=0.59), BMI (rg=0.20), age of first birth (rg=-0.26), and years of schooling (rg=-0.17) independently correlated with CWP (supplementary figure S5B, supplementary table S6).

### Tissue-specific expression of CWP mapped gene-sets

The results of functional annotation of GWAS significant loci are presented in supplementary figure S6 (supplementary results). Four different gene mapping strategies were implemented in FUMA (genome-wide gene-based association analysis, positional, eQTL, and chromatin interaction mapping) linking annotated SNPs to 89 genes of which 9 *(MST1, GMPPB, APEH, RNF123, ARVCF, AMIGO3, IP6K1, TANGO2*, and *TRAIP)* were identified using all four methods (figure 5A,B)[33]. At genomic risk loci 3p21.31, 3q22.1, and 22q11.21 a total of 54, 10 and 3 genes were prioritized by either eQTL or chromatin interaction mapping, respectively (figure 5C,D). Mapped genes were investigated for tissue-specific gene expression and for gene-set enrichment. In 54 specific GTEx tissues types, differentially expressed gene sets were enriched for skeletal muscle, several brain tissues, heart, whole blood, pancreas, and transverse colon (figure 6A, supplementary table S7). In 30 general GTEx tissue types, differentially expressed gene sets enriched for skeletal muscle, pancreas, heart, blood and brain (figure 6B, supplementary table S8). In both sets of GTEx tissues, overall enrichment for differentially expressed gene sets containing *RNF123* and *ATP2C1* gene were stronger for skeletal muscle than other tissues. *RNF123* was found to be highly expressed in skeletal muscle compared to other tissue types (figure 6C). None of the hallmark gene sets available in the molecular signature database was identified in the analysis.

**Figure 5.**
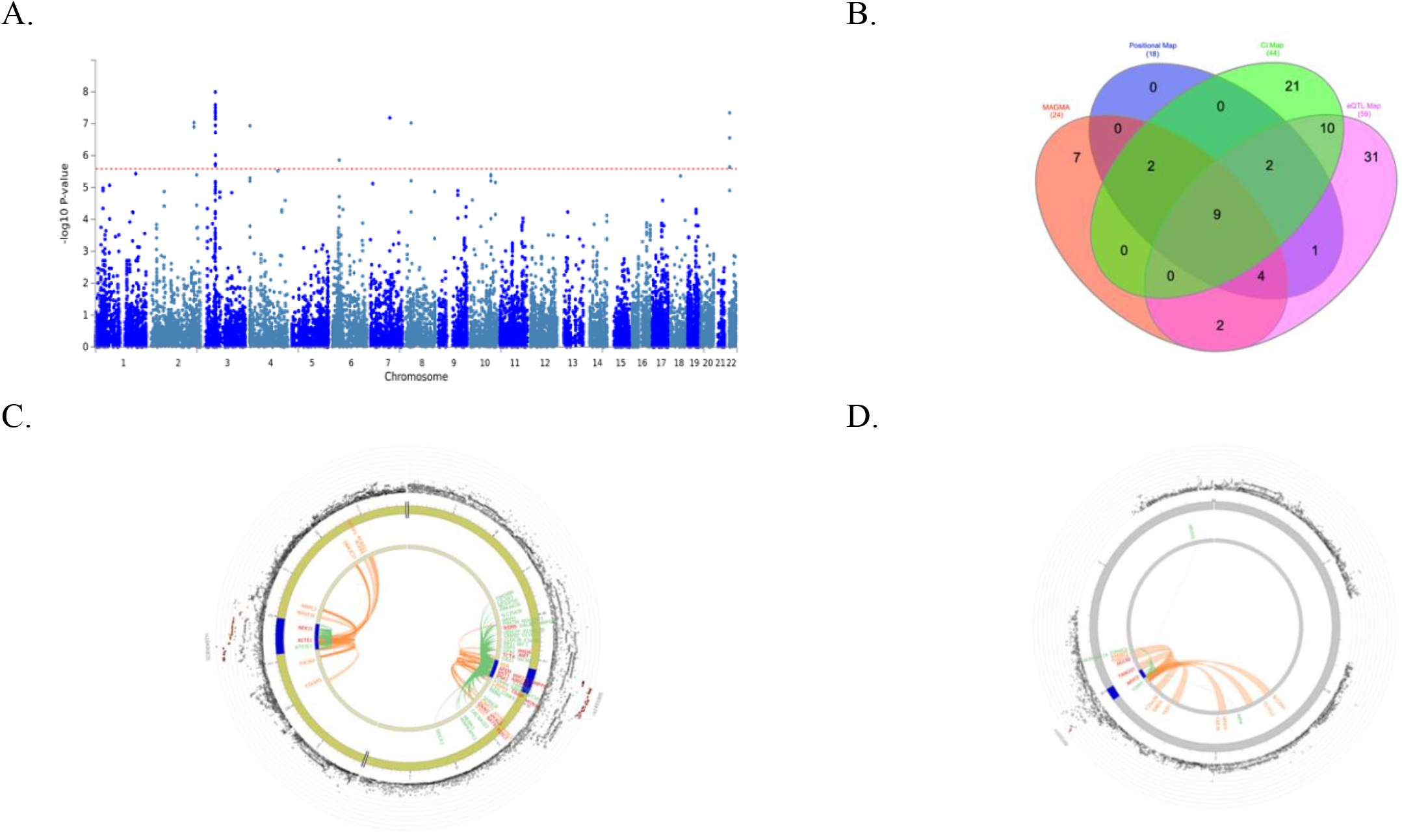
(A) Manhattan plot of the genome-wide gene-based association analysis, (B) Venn diagram showing overlap of genes implicated by genome-wide gene-based analysis implemented in MAGMA, positional mapping, chromatin interaction mapping (Ci Map), and eQTL mapping, (C) & (D) The circus plot displaying chromatin interactions and eQTLs on chromosomes 3 and chromosomes 22, respectively. The y-axis shows the ─log10 transformed two-tailed p-value of each gene from a linear model and the chromosomal position on the x-axis. The red dotted line indicates the Bonferroni-corrected threshold for genome-wide significance of the gene-based test. The most outer layer of the circus plot displaying Manhattan plot with –log10 P-values for CWP associated SNPs. Each independent SNP is presented with RSID. LD relationship between independent SNPs at the locus and their proxies are indicated with red (r 2 > 0.8) and orange (r 2 > 0.6). Grey SNPs indicate minimal LD with r2 ≤0.20. The outer circle represents chromosome with genomic risk loci are highlighted in blue. Either chromatin interaction or eQTLs mapped genes are displayed on the inner circle. Chromatin interactions and eQTLs mapped genes are presented in orange or green color, respectively. Genes mapped with both approaches are colored red.

**Figure 6.**
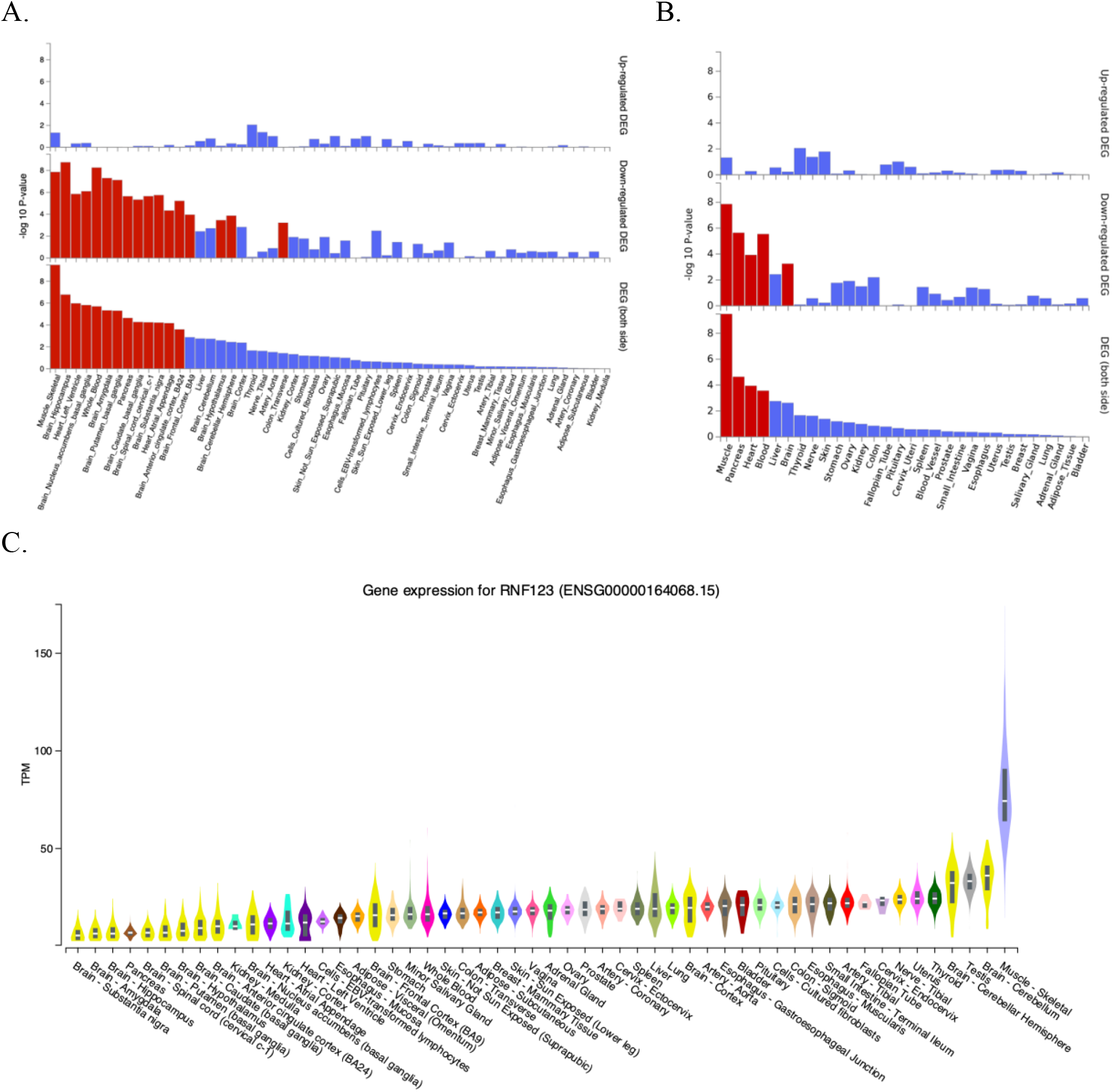
(A) Differentially expressed gene (DEG) plots for CWP in 54 tissue types from GTEX v8, (B) DEG plots for CWP in 30 general tissue types from GTEX v8 and (C) Differential expression of *RNF123* gene across tissue types from GTEX v8 (A, B) In both plots, the y-axis represents the ─log10 transformed two-tailed P-value of the hypergeometric test. Significantly enriched DEG sets (Bonferroni corrected p-value < 0.05) are highlighted in red. (C) Y-axis represents TPM (transcripts Per Million) and X-axis represents the GTEx (Version-8) tissues. The figure was adapted from GTEx Portal (https://www.gtexportal.org/home/gene/ENSG00000164068).

### Colocalization with skeletal muscle and dorsal root ganglion eQTLs

Colocalization analysis identified 93% of probability of shared eQTL variant *rs6809879* which controls *CDHR4* expression in the skeletal muscle and CWP association signal near *RNF123* locus (supplementary table S9, supplementary figure S7A). Additionally, significant colocalization was found for *rs13093525*, which controls *APEH* expression in DRG at exon-level (72% of probability of shared variant with *RNF123* locus). Both *rs6809879* and *rs13093525* were in complete LD with lead SNP *rs1491985(R2=1)* (supplementary table S10, supplementary figure S7B). No evidence of skeletal muscle or DRG eQTL colocalization was observed for *ATP2C1* and *COMT* loci.

## Discussion

CWP is a highly prevalent condition with moderate heritability and serves as a cardinal diagnostic feature of fibromyalgia. Therefore, our findings are of importance for better understanding the genetic basis of fibromyalgia. We report here the largest GWAS of CWP to date using 249,843 participants from the UKB, identifying 3 genome-wide significant loci implicating *RNF123, ATP2C1*, and *COMT* genes. The association in *RNF123* was replicated, whereas *ATP2C1* showed a suggestive association, and the *COMT* locus did not replicate in 41,296 individuals from independent cohorts.

*RNF123* (Ring Finger Protein, also known as KPC1) gene encodes E3 ubiquitin-protein ligase, which has a role in cell cycle progression, metabolism of proteins and innate immunity[34, 35]. Gene expression was higher in skeletal muscle than other tissues. It is not clear how *RNF123* contributes to CWP but it does have a role in malignancy[36, 37], osteoclast differentiation[38] and depression[39]. Consistent with this, an increased cancer incidence and poor cancer survival[40], poor bone health[41, 42], and co-existence of depression[43] have been reported in patients with fibromyalgia suggesting a shared pathway between these conditions and CWP. However, colocalization of *RNF123* locus with skeletal muscle, and DRG eQTLs reiterates the functional relevance of this locus in CWP and is in keeping with the symptomatic skeletal muscle tenderness of which patients complain.

*ATP2C1* (ATPase secretory pathway Ca2+ transporting 1) demonstrated a suggestive association in replication (p=0.0227). There was a consistent direction of effect for *ATP2C1* locus in all six replication cohorts except for ELSA, where we used a proxy SNP, which had close to zero effect size (beta= −0.0004±0.0110). Both positional and eQTL gene mapping strategies implicated *ATP2C1* with CWP. The *ATP2C1* gene encodes for the adenosine triphosphate (ATP)-powered magnesium-dependent calcium pump protein hSPCA1, which mediates Golgi uptake of cytosolic Ca(2+) and Mg(2+)[44]. A loss of function mutation in the *ATP2C1* leads to autosomal dominant Hailey-Hailey disease (HHD), a skin condition characterized by blistering and erosion of the epidermis[45]. Interestingly, published evidence shows HHD can be treated successfully with low-dose naltrexone (LDN), an opioid receptor antagonist, which has been used in the management of fibromyalgia, although opioids are now not recommended[46, 47]. Additionally, the role of calcium regulation in pain processing is well known[48-50]. Pregabalin is an α2δ calcium channel antagonist, which is currently used in the management of fibromyalgia[51], all supporting the regulation of calcium influencing CWP/fibromyalgia. This is the first study to implicate *ATP2C1* with musculoskeletal pain using an agnostic approach.

*COMT* is one of the most studied genes in human pain[52]. Almost 30 SNPs and three haploblocks of the *COMT* gene have been studied in acute clinical, experimental and chronic pain. *Rs4680* of the *COMT* gene is extensively studied in many pain phenotypes such as pain sensitivity, TMD, and fibromyalgia[53]. Across multiple ethnic populations, *rs4680* was implicated in the development of fibromyalgia[14]. However, a meta-analysis of 8 case-control studies (589 fibromyalgia cases and 527 controls) did not confirm earlier association[54]. To date, the largest study assessed the association between *COMT* haplotypes (*rs4680, rs4818, rs4633*, and *rs6269*), and fibromyalgia included 60,367 participants (2,713 ICD-9 diagnosed fibromyalgia) and found no association[55]. They have also been refuted in other European CWP samples[21, 56] and a large candidate gene study of fibromyalgia[57]. However, we identified *rs165599*, located at 3’UTR of *COMT*, associated with CWP in the discovery sample but not in the meta-analysis or any of the six replication cohorts. This variant is not in LD with previously studied *COMT* SNPs *rs4680, rs4818, rs4633* and *rs6269*, and was found not to be associated with chronic musculoskeletal pain including CWP neither when studied as single SNP nor as a part of a haploblock[58-60]. Several explanations of our non-replication of *COMT* locus are possible. First, there was lower power pertaining to overall meta-analysis, which was estimated at 48%, based on the effect size observed in the discovery sample (n=249,843), replication sample size (n=43,080), and the number of tests conducted (n=3). Our meta-analysis did have 90% power to detect a relative risk as small as 1.04 but the estimated *COMT* effect was only 1.012 (beta=0.0027±0.004; OR=1.012, 95% CI=0.97–1.05). Our replication sample size was, however, larger than many of the earlier studies (included 122 to 3811 participants) that reported the association between *COMT* and CWP[20, 61]. Second, we observed a tendency towards non-significance for the *COMT* locus in the sensitivity GWAS due to the exclusion of participants with non-musculoskeletal pain from the control group suggesting that *COMT* predisposes to chronic pain in general. Finally, genetic factors underlying chronic pain and psychiatric co-morbidity (e.g., depression and neuroticism) are known to be shared[62]. However, previous GWAS on chronic pain[29, 63, 64], depression[65] and neuroticism[66] have failed to detect an association with *COMT*. Thus, if there is a role of *COMT* in CWP, it is likely minimal.

Epidemiological studies have consistently reported higher BMI to be associated with an increased risk of CWP[10-12]. Our analysis showed significantly higher BMI in CWP cases compared to controls (p<0.0001) in all cohorts except RS-3. In line with this, we observed a positive genetic overlap between BMI and CWP independent of genetic confounders. Similarly, genetically independent pairwise genetic correlation for depressive symptoms, age of first birth, and years of schooling was seen with CWP. These findings indicate the presence of shared molecular pathways underlying these traits.

Functional analysis showed that prioritised genes are differentially expressed in skeletal muscle, several areas of the CNS, pancreas, whole blood and heart tissues. These findings suggest the involvement of nervous, musculoskeletal, and neuroendocrine systems in CWP. These physiological systems have been implicated in fibromyalgia in previous studies[67-69]. Evidence suggests that both peripheral and central pain mechanisms influence CWP[70, 71]. We observed overall stronger enrichment for differentially expressed gene sets in skeletal muscle than in other GTEx tissues such as brain. Also, skeletal muscle and DRG eQTLs colocalize with the *RNF123* locus. These findings suggest a substantial involvement of peripheral pain mechanisms in CWP. However, we established tissue specificity in the expression of multiple genes likely related to CWP. An in-depth investigation of these genes may provide additional insight into the pathogenesis of CWP.

There are several limitations to the study. The case definition of CWP depends on self-report together with exclusion of other conditions with symptoms leading to chronic pain[72]. A clinical diagnosis of CWP would have been infeasible in a sample this large. Also, we used common SNPs to estimate the heritability of CWP, so the contribution of other variants in the heritability estimated remains unknown. Other musculoskeletal pain assessed in UKB (knee, hip, back, and neck or shoulder pain) exhibited SNP-heritability on the liability scale in the range 0.08–0.12[62], and these estimates are considerably lower than our estimate for CWP, suggesting CWP is a trait of high genetic influence.

In summary, this study identified a novel association for CWP in the gene *RNF123* and suggested the role of calcium regulation, by the involvement of the *ATP2C1* locus. The association of the *COMT* locus with CWP was not replicated, suggesting a small influence, if any. We found evidence that the epidemiological association of BMI and CWP is at least in part genetically mediated. Finally, our results suggest a profound role of peripheral mechanisms in the pathogenesis of CWP.

## Data Availability

Summary statistics from our GWAS discovery was deposited at Zenodo (https://doi.org/10.5281/zenodo.4298764). Other data relevant to the study are included in the article or uploaded as supplementary information.

## Competing interests

YSA is co-owner of Maatschap PolyOmica and PolyKnomics BV, private organizations, providing services, research and development in the field of computational and statistical, quantitative and computational (gen)omics.

## Contributors

MDSR prepared the manuscript. Discovery GWAS: MDSR, MF and FW designed the study. MDSR, YAT, MF and FW developed phenotype definition. MDSR performed data management and GWAS in the discovery cohorts. Replication: MDSR, MF and FW developed replication protocol and coordinated the study; MDSR, BW, JAW, MF and FW designed the study; BW, SB, KH, EF, KHveem, JAZ, HUNT All-in Pain authors, SOCC, JBJVM and FW collected replication cohort’s data; MDSR, BW, SB, KHveem, JAZ, SOCC and JBJVM were involved data management; MDSR, BW, SB, KH, EF, JAZ, HUNT All-in Pain authors, MF and FW developed phenotype definition; MDSR, BW, SB, SOCC and JBJVM analysed the data. MDSR performed replication meta-analysis. Post-GWAS bioinformatics: MDSR, YAT and SSh performed post-GWAS bioinformatics analysis. MF, YA and FW provided the statistical and bioinformatics consultation in the project. MDSR, KH, SOCC, YAT, SSh, YA, JBJVM, MF and FW interpreted the findings. FW was responsible for the overall supervision of the project. All authors commented on the manuscript and agreed on the final version to be published.

## Acknowledgements

We would like to thank all the participants of UK Biobank, The Nord-Trøndelag Health Survey, English Longitudinal Study of Aging, TwinsUK and Rotterdam study I, II and III. The contribution of inhabitants, general practitioners and pharmacists of the Ommoord district to the Rotterdam Study is gratefully acknowledged.

The UK Biobank study was approved by the National Health Service National Research Ethics Service (ref.11/NW/0382) and all participants provided written informed consent. Genome-wide association analysis was performed using the UK Biobank resource under project number 18219. The English Longitudinal Study of Ageing is jointly run by University College London, Institute for Fiscal Studies, University of Manchester and National Centre for Social Research. Genetic analyses have been carried out by UCL Genomics and funded by the Economic and Social Research Council and the National Institute on Aging. Data governance was provided by the METADAC data access committee, funded by ESRC, Welcome, and MRC. (2015-2018: Grant Number MR/N01104X/1 2018-2020: Grant Number ES/S008349/1). TwinsUK is funded by the Wellcome Trust, Medical Research Council, European Union, Chronic Disease Research Foundation (CDRF), Zoe Global Ltd and the National Institute for Health Research (NIHR)-funded BioResource, Clinical Research Facility and Biomedical Research Centre based at Guy’s and St Thomas’ NHS Foundation Trust in partnership with King’s College London. The Nord-Trøndelag Health Study (The HUNT Study) is a collaboration between HUNT Research Centre (Faculty of Medicine and Health Sciences, NTNU, Norwegian University of Science and Technology), Trøndelag County Council, Central Norway Regional Health Authority, and the Norwegian Institute of Public Health. The genotyping was financed by the National Institute of Health (NIH), University of Michigan, The Norwegian Research Council, and Central Norway Regional Health Authority and the Faculty of Medicine and Health Sciences, Norwegian University of Science and Technology (NTNU). The genotype quality control and imputation has been conducted by the K.G. Jebsen center for genetic epidemiology, Department of public health and nursing, Faculty of medicine and health sciences, Norwegian University of Science and Technology (NTNU). The Rotterdam Study is supported by the Erasmus MC University Medical Center and Erasmus University Rotterdam; The Netherlands Organization for Scientific Research (NWO); The Netherlands Organization for Health Research and Development (ZonMw); the Research Institute for Diseases in the Elderly (RIDE); The Netherlands Genomics Initiative (NGI); the Ministry of Education, Culture and Science; the Ministry of Health, Welfare and Sports; the European Commission (DG XII); and the Municipality of Rotterdam.

## Funding

MDSR received funding from the European Union’s Horizon 2020 research and innovation program IMforFUTURE, under H2020-MSCA-ITN grant agreement number 721815. The work of YAT was supported by PolyOmica and by the Russian Foundation for Basic Research (project 19-015-00151). The work of SZS was supported by the Federal Agency of Scientific Organizations via the Institute of Cytology and Genetics (project 0324-2019-0040-C-01/AAAA-A17-117092070032-4). The work of YSA was supported by PolyOmica.

## Patient and public involvement

Patients and/or the public were not involved in the design, or conduct, or reporting, or dissemination plans of this research.

## Patient consent for publication

Not required

## Ethics approval

Study-specific ethical approval was provided by the local ethics committees. Details provided in the supplementary material.

## Notes

### Competing Interest Statement

YA is co-owner of Maatschap PolyOmica and PolyKnomics BV, private organizations, providing services, research and development in the field of computational and statistical, quantitative and computational (gen)omics.

### Funding Statement

MDSR received funding from the European Unions Horizon 2020 research and innovation program IMforFUTURE, under H2020-MSCA-ITN grant agreement number 721815. The work of YAT was supported by PolyOmica and by the Russian Foundation for Basic Research (project 19-015-00151). The work of SZS was supported by the Federal Agency of Scientific Organizations via the Institute of Cytology and Genetics (project 0324-2019-0040-C-01/ АААА-А17-117092070032-4). The work of YSA was supported by PolyOmica.

### Summary of Updates

Author Yakov A. Tsepilov name was revised.

