## Supplementary Material for "Genome-wide association study identifies *RNF123* locus as associated with chronic widespread musculoskeletal pain"

Banner "**HUNT All-In Pain**"

Amy E Martinsen^1,2,3^, Anne Heidi Skogholt^1,4^, Ben Brumpton^1^, Cristen Willer^5^, Ingrid Heuch^2^, Ingunn Mundal^6^, Jonas Bille Nielsen^1,5,7^, Kjersti Storheim^8,9^, Kristian Bernhard Nilsen^10,11^, Lars Fritsche^12^, Laurent F. Thomas^1,4,13,14^, Linda M Pedersen^2^, Maiken E Gabrielsen^1^, Marianne Bakke Johnsen^1,3,8^, Marie Udnesseter Lie^3,8^, Oddgeir Holmen^15^, Synne Øien Stensland^8,16^ , Wei Zhou^17,18^

Affiliations:

1 K. G. Jebsen Center for Genetic Epidemiology, Department of Public Health and Nursing, Faculty of Medicine and Health Sciences, Norwegian University of Science and Technology, Trondheim, Norway.
2 Department of Research, Innovation and Education, Division of Clinical Neuroscience, Oslo University Hospital, Oslo, Norway.
3 Institute of Clinical Medicine, Faculty of Medicine, University of Oslo, Oslo, Norway.
4 Department of Clinical and Molecular Medicine, Norwegian University of Science and Technology, Trondheim, Norway.
5 Department of Internal Medicine, Division of Cardiovascular Medicine, University of Michigan, Ann Arbor, 48109, MI, USA.
6 Department of Health Science, Molde University College, Molde, Norway.
7 Department of Epidemiology Research, Statens Serum Institut, Copenhagen, Denmark.
8 Research and Communication Unit for Musculoskeletal Health (FORMI), Department of Research, Innovation and Education, Division of Clinical Neuroscience, Oslo University Hospital, Oslo, Norway.
9 Faculty of Health Sciences, Department of physiotherapy, Oslo Metropolitan University, Oslo, Norway.
10 Department of Neuromedicine and Movement Science, Faculty of Medicine and Health Sciences, Norwegian University of Science and Technology (NTNU), Trondheim, Norway.
11 Department of Neurology, Oslo University Hospital, Oslo, Norway.
12 Center for Statistical Genetics, Department of Biostatistics, University of Michigan, Ann Arbor, 48109, MI, USA.
13 BioCore - Bioinformatics Core Facility, Norwegian University of Science and Technology, Trondheim. Norway.
14 Clinic of Laboratory Medicine, St.Olavs Hospital, Trondheim University Hospital, Trondheim, Norway.
15 HUNT Research Center, Department of Public Health and Nursing, Faculty of Medicine and Health Sciences, Norwegian University of Science and Technology, Trondheim, Norway.
16 NKVTS, Norwegian Centre for Violence and Traumatic Stress Studies.
17 Department of Computational Medicine and Bioinformatics, University of Michigan, Ann Arbor, MI, USA.
18 Analytic and Translational Genetics Unit, Massachusetts General Hospital, Boston, Massachusetts, USA.

| Supplementary Methods |  |
| --- | --- |
| Supplementary Results |  |
| Supplementary Figure S1 | Study Flowchart of cases and controls |
| Supplementary Figure S2 | The QQ plot of GWAS summary statistics of CWP |
| Supplementary Figure S3 | Manhattan plot of sensitivity GWAS of CWP |
| Supplementary Figure S4 | Heatmap of genetic correlations for 23 complex traits and CWP |
| Supplementary Figure S5 | Hierarchical clustering of genetic correlations and heatmap of partial genetic correlations. |
| Supplementary Figure S6 | Functional consequences of candidate SNPs in genomic risk loci annotated by ANNOVAR |
| Supplementary Figure S7 | Colocalization of CWP associated locus (*RNF123*) with skeletal muscle and dorsal root ganglion eQTL |
| Supplementary Table S1 | Genotyping and imputation methods across all cohorts. |
| Supplementary Table S2 | Lead SNPs significantly associated with CWP in UK Biobank |
| Supplementary Table S3 | Replication findings for reach cohorts |
| Supplementary Table S4 | Sample-size based meta-analysis findings of replication SNPs |
| Supplementary Table S5 | Standard error based meta-analysis findings of replication SNPs |
| Supplementary Table S6 | Genetic correlations and partial genetic correlations between seven complex traits and chronic widespread pain |
| Supplementary Table S7 | Differential gene set enrichment in 54 specific tissue types from GTEx |
| Supplementary Table S8 | Differential gene set enrichment in 30 general tissue types from GTEx |
| Supplementary Table S9 | Colocalization of *RNF123* locus with muscle skeletal eQTL signals |
| Supplementary Table S10 | Colocalization of *RNF123* locus with DRG eQTL signals at exon-level |
| References | |

**Supplementary Methods**

**Description of study cohorts**

**UKB.** UKB is a population cohort comprising 502,682 individuals aged 40-73 years at recruitment, who are registered with a general practitioner within the UK National Health Service. Around 9.2 million individuals living within 25 miles of UKB assessment centre (n=22) located in England, Scotland, and Wales were invited to take part in the study between 2006 and 2010. Data collected was primarily self-reported. Participants were provided with touchscreen computer-based questionnaire and also attended a face-to-face interview administered by trained nurses. Each participant provided phenotypic and health-related information (e.g. pain, lifestyle and environmental) and biological samples (e.g. blood, urine and saliva). Following the Declaration of Helsinki, written informed consent was obtained from each participant[1]. UK Biobank’s study protocol is available publicly (<http://www.ukbiobank.ac.uk/wp-content/uploads/2011/11/UK-Biobank-Protocol.pdf?phpMyAdmin=trmKQlYdjjnQIgJ%2CfAzikMhEnx6>) and research activities were reviewed and approved by the North West Research Ethics Committee (REC reference no. 16/NW/0274). Majority of cohort participants (94%) self-reported to be white ancestry[2].

**HUNT.** The Nord-Trøndelag Health Study (HUNT) (<https://www.ntnu.edu/hunt>) is a population based, longitudinal study carried out in Nord-Trøndelag county in Norway. It comprises an ethnically homogenous, primarily Caucasian population. The study has been carried out in several waves (HUNT1-4), and in each survey, all inhabitants aged ≥ 20 years were invited to participate. A range of health-related data were obtained, both through questionnaires and clinical examinations. DNA from whole blood was collected in HUNT2 (1995-1997) and HUNT3 (2006-2008), with genotypes being available for 71,860 participants. Both surveys also included questions to define CWP[3]. A more detailed description of the HUNT Study is available elsewhere[4]. All study participants provided an informed, written consent to use their data and biological samples for research, and the study was approved by the Regional Committee of Medical and Health Research Ethics in Norway (REK #2015/573).

**TwinsUK.** TwinsUK cohort ([www.twinsuk.ac.uk](http://www.twinsuk.ac.uk)) comprises approximately 13,000 MZ and DZ twins aged between 18 to 93 living in the United Kingdom. TwinsUK registry commenced in 1992 and in later years additional twins were recruited to understand heritability, the genetic architecture of common diseases and the healthy ageing process. Participants of the TwinsUK cohort are predominantly females. Detailed phenotypic and omics data were collected from twins. All participants were recruited following the Declaration of Helsinki, and all research projects were approved by the Research Ethics Committee of the St. Thomas’ Hospital. All participants of TwinsUK registry provided written consent. Information on CWP and other omics are available from the TwinsUK participants[5]. This study includes participants who responded to CWP questionnaire between 2002–2013.

**RS.** RS ([www.epib.nl/research/ergo](http://www.epib.nl/research/ergo)) is a population-based prospective cohort study in the district of Rotterdam, the Netherlands and comprised of three independent cohorts. The first cohort started in the 2^nd^ half of 1989 with 7,983 persons aged ≥ 55 to 106 years living in Ommoord district in the city of Rotterdam called Rotterdam Study I (or RS-I). In the second cohort (Rotterdam Study II (or RS-II)), 3011 participants aged 55 in the year 1999 were added to the study. In the third cohort (Rotterdam Study III (RS-III)), 3932 participants aged between 45–54 years were added in the study. All three RS study participants were interviewed for 2 hours at home and extensively examined (e.g., imaging heart, blood vessels, eyes, skeleton and brain) for 5 hours in a research facility which was repeated in every 3 to 4 years in a research facility. Biospecimens were collected during the research facility visit. Informed consent was obtained from each participant, and the medical ethics committee of the Erasmus Medical Centre Rotterdam approved the study[6, 7].

**ELSA.** ELSA (<https://www.elsa-project.ac.uk/>) is a prospective open cohort comprised of a representative ageing population of England. This study was designed to capture the experience of the aged population in the 21^st^ century. The study is ongoing but has collected a wide range of high-quality data in the last two decades, which includes health, economic, social, psychological, cognitive, biological and genetic data. At present, the ELSA study had completed eight waves (w-1 to w-8) of data collection between 2002-2017. In each wave, data was collected via computer-assisted personal interview, a self-reported questionnaire, tests for cognitive function and walking speed. The nurse collected biological samples from participants. In the computer-assisted personal interview, along with other modules (e.g., household demographics, individual demographics, work and pensions), a health module was administered to all respondents which covered long-standing illness or disability, eyesight and hearing, specific diagnoses and symptoms, and pain etc. Ethical permission for all the ELSA waves was provided by the National Research Ethics Service (MREC/01/2/91)[8]. Use of ELSA data for this project was approved by METADAC data access committee (application reference: MDAC-2019-0928-03A-FREYDIN).

**Phenotype definition – Discovery cohort**

**UKB.** The UKB participants were provided with a touchscreen questionnaire and asked “In the last month have you experienced any of the following that interfered with your usual activities? Possible answers to choose from were ‘none of the above’; ‘prefer not to answer’; pain at seven different body sites (head, face, neck/shoulder, back, stomach/abdomen, hip, knee); or ‘all over the body’. Unless reported pain “all over the body”, participants could report more than one pain site. Those reported to have pain in the last month were further asked if the pain lasted for 3+ months. Participants with three months of “pain all over the body” were considered as cases of CWP (n=5,440). Also, those reporting simultaneous pain in knee, shoulder, hip and back lasting for 3+ months were considered as cases (n=2,132). In addition, we used data field 20002 (Non-cancer illness) where participants either self-reported “fibromyalgia” or described the condition to the interviewee who provided the diagnosis. A total of 726 participants reported a diagnosis of fibromyalgia which was added to the study cases.

**Phenotype definition – Replication cohorts**

**HUNT.** The definition of CWP used in this study was published before [9]. In brief, participants were asked the screening question “Have you during the last year continuously for at least 3-months had pain and/or stiffness in muscles and joints?”. Those who replied “yes” were requested to mark the location of nine pain sites (neck, shoulders, elbows, wrist/hands, upper back, low back, hips, knees, and/or ankles/feet). These nine anatomical pain sites were taken from the Nordic Questionnaire[10], and have been shown to be reliable in estimating low back and upper limb, and neck symptoms during the past year[11]. CWP cases were defined as those with pain located in the axial skeleton (neck, upper back, or lower back), above the waist (neck, shoulders, elbows, wrist/hands, or upper back), and below the waist (lower back, hips, knees, or ankles/feet). In HUNT3 cases were also required to have bilateral presence of the pain, but not in HUNT2, where no question on laterality was included. Controls were defined as participants who were free from any form of chronic musculoskeletal pain (< 3 months) in HUNT-2 and HUNT-3. Based on International Statistical Classification of Diseases (ICD)-10 codes participants with a diagnosis of rheumatoid arthritis, polymyalgia rheumatica, arthritis not otherwise specified (NOS), systemic lupus erythematosus, ankylosing spondylitis were excluded from the study. The final sample included in the replication analysis consisted of 10,556 CWP cases and 13,239 controls.

**ELSA.** Study participants were asked about their experience of pain using computer-assisted personal interview. Pain questions asked differed in their contents between the waves. In all waves, participants were asked “are you often troubled with pain?”, following a “yes” response follow up questions were asked to identify the number of pain sites and severity and/or duration of pain. The methods of ascertaining pain sites differed between waves. In waves 1 and 2, participants were asked to report pain in 4 musculoskeletal sites (back, hip, knee and feet) on a scale of 0 (no pain) to 10 (severe excruciating pain). In contrast, in waves 3 to 8, participants were asked to report their experience of pain in the 7 sites (back pain, hip pain, knee pain, feet pain, mouth pain, pain elsewhere and pain all over) and they could choose as many options as they liked. The severity of pain was asked in all waves, and duration of pain was requested explicitly in waves 4, 5 and 6. In the study, we defined CWP if participants reported pain all over or simultaneous pain in the back, hip and knee or back, hip, and feet or back, knee and feet which was lasting for more than three months or in their severity as moderate to severe (in the absence of pain duration). We defined controls if participants reported “No” to the question “are you often troubled with pain?”. Finally, we made a composite CWP binary variable by merging all cohorts where CWP cases identified in all waves served as cases. In contrast, controls were those found to be controls in any waves but never became cases in the earlier or later waves. A total of 1,679 cases and 5,304 controls with genotype data were included in the replication analysis.

**TwinsUK.** CWP information was collected on five occasions using questionnaires between 2002-2014. On three instances London Fibromyalgia Epidemiology Study Screening Questionnaire (LFESSQ)[12] was administered. In the other two instances broader information was collecting including site-specific questions or a mannequin was provided to report pain sites. Based on information collected, we defined CWP in the study as pain in the middle, left and right side of the body, above and below diaphragm lasting for three months or more[12]. A composite CWP variable was made by merging all five data collection time where cases were those ever reported to have CWP and controls were who did not fulfil the criteria of CWP in any of the waves. Participants with inflammatory diseases (n=67) and missing zygosity were excluded from the study. Participants who reported disabled low-back pain (n=455) were excluded from the controls. Finally, 1111 cases and 3556 controls with genotype data were included in the replication analysis.

**RS.** The RS study participants reported painful body sites (pain during at least half of the days during the last six weeks) using a pain homunculus. CWP was defined if participants reported pain sites in the left side of the body, in the right side of the body, above and below the waist, and in the axial skeleton. Same CWP definition was used in previous GWAS[7]. Controls were those reported no pain or any form of chronic musculoskeletal pain (≥ 3 months).

##### **Selection of proxy SNPs in ELSA cohort**

To identify proxy SNPs, we looked for ELSA genotyped SNPs around ±250kb of the discovery SNPs. We choose proxies for replication analysis using criteria that the SNP had minor allele frequency closer to the SNP identified in the discovery, showing highest R^2^ (> 0.80) with the discovered SNP and had lowest genetic association p-value in the discovery cohort. The *rs9870858* and *rs1732984*8 have been considered as best proxy SNPs for *rs1491985* and *rs10490825*, respectively.

### **Statistical analysis and in-silico follow-up**

**Discovery association analysis**

We applied the following filters to discovery analysis: minor allele frequency (MAF) ≥ 0.01, imputation quality scores (INFO) ≥ 0.70, SNPs and individuals missingness rates not exceeding 0.02. Plink V.2[13] has been used to determine SNPs passing Hardy-Weinberg equilibrium (HWE) threshold p > 1E-06. P-value threshold 5E-08 was used to declare GWAS significance.

#### **Identification of independent SNPs**

To identify independent SNPs located in GWAS significant loci (p<5E-08) we used multi-SNP-based conditional & joint association (COJO)[14] analysis implemented in software package GCTA[15]. A stepwise model selection procedure was used to identify independently associated SNP by conditioning on other significant SNPs at the locus. SNPs with minor allele frequency ≤ 0.01 was excluded. Randomly selected 50,000 European ancestry participants from the UK Biobank were used as LD reference sample for the COJO analysis. In addition to COJO, we used Functional Mapping and Annotation of genetic associations (FUMA) v1.3.4[16] to identify independent SNPs at p<5E-08 by examining the relationship between independent SNPs at r2 < 0.1. The 1000 genome phase-3 European ancestry data was used as a reference panel to define LD blocks (<250 kb apart, MAF ≥ 0.01). Findings of GCTA-COJO and FUMA were identical.

**Replication and Meta-analysis**

**HUNT Association testing.** We performed association testing between independent SNPs and CWP using the Scalable and Accurate Implementation of Generalized mixed model (SAIGE)[17], which uses a generalized mixed model to account for sample relatedness and cryptic population structure. We performed a mixed-effects linear regression model, including age, sex, genotype batch, and the first four genetic principal components as covariates.

**TwinsUK association testing.** We performed a linear mixed-effects model using Genome-wide Efficient Mixed Model Association (GEMMA) v0.98.1[18] to estimate the effect of each independent SNP. Regression models were adjusted for age, sex, and the genetic relatedness matrix.

**ELSA association testing.** We performed a mixed-effects linear model using Genome-wide Complex Trait Analysis (GCTA) v1.91.7 beta1[15] to estimate the effect of each independent SNPs. Regression models were adjusted for age, sex, and the genetic relationship matrix.

**RS association testing.** For all three RS cohorts, we performed a linear regression model using PLINK v1.9.[13] to estimate the effect of each independent SNPs. Assuming homogenous study population, RS cohorts were adjusted for age and sex only.

**Meta-analysis of replication cohorts.** Association findings of each SNP across all replication cohorts were meta-analysed using fixed effects model with sample size and inverse-variance weighting implemented in METAL[19]. Between-study heterogeneity was assessed using I^2^ statistics. Multiple testing correction was applied to declare significance following meta-analysis (0.05/3= 0.017). We performed both sample size and standard error based meta-analysis. Power calculation showed that replication meta-analysis power for three independent SNPs ranges between 46.3 to 49.7%.

#### **Genomic inflation, heritability, genetic correlation and partial genetic correlations**

LD score regression (LDSR)[20] was used to assess inflation (λ_GC_) in test statistics and to distinguish confounding from polygenicity. We also used LDSR to estimate SNP-based heritability of CWP, which was converted on the liability scale. We measured the genetic correlation (GC) between CWP and non-UKB complex traits from LD-hub[21] using LDSR tools[20]. A total of 209 complex traits were used for GC analysis. Precomputed LD scores using 1000 Genomes European data restricted to HapMap3 SNPs (n=1,217,311) were used to calculate both SNP heritability and genetic correlations. Precomputed LD scores and the list of HapMap3 SNPs were obtained from <https://data.broadinstitute.org/alkesgroup/LDSCORE/>.

Bonferroni-corrected p-value < 0.01/209 = 4.78E-05 was used to declare significance for GC analysis. Based on hierarchical clustering, we identified 7 clusters of genetically correlated traits, of which seven representative traits were chosen for partial GC analysis. Partial GC quantifies the proportion of GC, which is not influenced by other traits. Visualization of GC, hierarchical clustering and partial GC implemented in R using package "corrplot" with basic "hclust" function. Bonferroni-corrected p-value < 0.01/7 = 0.001 was used to declare significance for partial GC analysis.

#### **Functional annotation**

To identify the functional importance of GWAS loci at p <5E-08, we used ANNOVAR[22] implemented in FUMA[16]. Independent SNPs identified at r^2^<0.6 within a 250kb window and their LD proxies with MAF ≥ 1% were selected using 1000 Genomes Project Phase 3 as a reference panel. All independent SNPs and proxy SNPs were taken forward for annotation in ANNOVAR with Ensembl genes build v92. Additionally, CADD score (a score >12.37 considered to be pathogenic), RegulomeDB (RDB) scores (which ranges from 1 to 7 where the lower score indicates a higher likelihood of having a regulatory function), and 15-core chromatin states (chromatin state <8 indicates an open chromatin region with higher accessibility as the score decreases) were annotated. All these features were embedded in the FUMA web tool.

**Gene mapping**

We used four different strategies (genome-wide gene-based association analysis, positional, eQTL, and chromatin interaction mapping) for gene mapping. MAGMA (Multi-marker Analysis of GenoMic Annotation) v1.07[23] was used for gene-based genome-wide association analysis (GWGAS), which was implemented in a web tool FUMA[16] v1.3.6. In GWGAS analysis, SNPs from the CWP GWAS summary statistics were mapped to 19261 protein-coding genes using gene definition of NCBI Build 37/UCSC hg19. All SNPs locating within ±50Kb of the gene body were used to calculate a gene test-statistics (p-value) using default SNP-wide mean model. The major histocompatibility complex (MHC) region was excluded from the analysis. For the calculation of LD 1000 genome phase-3 European ancestry data was used as a reference. Results were presented with Bonferroni correction to control for multiple testing (P < 0.05/19,261= 2.6E-6).

For the positional gene mapping, ANNOVAR annotated SNPs were mapped to protein-coding genes within 10kb window from the human reference assembly (GRCh37/hg19) using FUMA. For the eQTL mapping, all independent SNPs and proxy SNPs identified by FUMA were mapped to all eQTL data repositories available in the FUMA with default settings. All SNPs were mapped to genes where the allelic variation of SNP affects the expression level of those genes up to 1 Mb. An FDR (false discovery rate) threshold < 0.05 was used to define significant eQTL association. In the chromatin interaction mapping, all candidate SNPs were mapped to genes’ promoter regions (defined with a window of 250bp upstream and 500bp downstream of TSS) based on significant chromatin interaction. This mapping strategy does not require distance boundary; therefore, genes located in long-distance can be mapped. When an independent SNP is located in a region interacting with a region containing several genes, then all of those genes were mapped with that SNP. We used Hi-c data of 21 tissues and cell types from GSE87112 available in FUMA by default for chromatin interaction mapping. To prioritise candidate genes, we performed the filtering of candidate SNPs overlapping with enhancers and promoters predicted from 111 tissue/cell types from the Roadmap Epigenomics Project. This strategy reduces gene number and increases the likelihood that the remaining genes are biologically relevant. An FDR <1E-06 were used to detect significant interaction.

#### **Tissue specificity and gene-set enrichment analyses**

Tissue and gene set enrichment analyses were conducted with GENE2FUNC, an integrated process of FUMA[16] web tool. A total of 89 mapped genes identified by GWGAS, positional, eQTL or chromatin interaction mapping were used as input*.* Tissue specificity for 54 specific tissues and 30 general tissues obtained from the GenotypeTissue Expression (GTEx) v8 database were tested using previously defined differentially expressed gene (DEG) sets. All mapped genes were tested against each DEG sets with the hypergeometric test. Additionally, an overrepresentation of mapped genes in any of the well-defined hallmark gene sets available in the molecular signature database (MsigDB) were tested. Tissue specificity and gene-set enrichment were conducted using FDR adjusted p-value threshold <0.05 and minimum overlapping genes with gene-sets ≥2. All genes available by default were used as background gene-set for the enrichment analysis. All of these analyses were performed, excluding the MHC genomic region.

**Colocalization analyses using skeletal muscle and dorsal root ganglia eQTLs**

We aimed to explore the cis-regulation of CWP associated variants in both skeletal muscle (n=706) and human DRG (n=214) using publicly available eQTL data (skeletal muscle: <https://gtexportal.org/home/>; DRG: <http://diatchenko.lab.mcgill.ca/DRG-eQTLs/>). SNPs regulating the expression level of a gene known as eQTLs. Cis-acting eQTLs were located within ≤1Mb of the transcription start site of the target gene [24]. Details of skeletal muscle and DRG eQTLs are available here[25, 26]. We extracted the summary statistics of SNPs associated with CWP at 1E-05 and located within a 200-kb window around GWAS independent SNPs. Extracted SNPs overlapping with skeletal muscle and DRG eQTLs were used for colocalization. Before colocalization analysis with GTEx skeletal muscle eQTLs, CWP-GWAS associated RSIDs were aligned to the human reference genome build GRCh38 using LiftOver tool (<https://genome.ucsc.edu/cgi-bin/hgLiftOver>). We applied Bayesian colocalization method (coloc)[27] with CWP prevalence and “cc” trait type as parameters to integrate CWP-GWAS with skeletal muscle and DRG cis-eQTLs data assuming a single causal variant underlying the locus. Colocalization of skeletal muscle eQTLs was assessed at gene-level. For DRG, both gene- and exon-level cis-eQTLs were assessed for colocalization. A total of five hypotheses were tested to evaluate colocalization, H0: there is no causal variant for both traits); H1 or H2: causal variant associated with either trait-1 or trait-2, H3: two independent causal variants for trait-1 and trait-2; and H4: one single causal variant associated with both traits. Coloc generates higher posterior probability (PP) to test each hypothesis. A higher posterior probability for H3 (PP3) supports the presence of two independent variants for both traits. A higher posterior probability for H4 (PP4) supports the presence of single independent variants affecting both traits. We reported eQTL SNP at the locus having lowest p-value as evidence of colocalization.

**Supplementary Results**

#### **Functional annotation using ANNOVAR**

Independent SNPs and their proxies were annotated for functional consequences for using ANNOVAR. In total, 225 candidate SNPs were used for annotation. The results of ANNOVAR annotation presented in supplementary table S6 and supplementary figure S6. Majority of the annotated SNPs were intronic (83.6%). None of the annotated SNPs were non-synonymous. We found 5 synonymous variants located in genes *ATP2C1* *(rs16835513), BSN (rs4855885), MST1 (rs3020779), TRAIP (rs35129566)* and *ARVCF (rs2073747)*. A total of 4% (n=9) of annotated SNPs having CADD score >12.37 indicating deleterious nature of these SNPs, of which three SNPs *(rs62280752, rs28362548* and *rs62282192)* were located at gene *ATP2C1*. An RDB score <2 was observed for 9.3% (n=21) of the SNPs indicate that these variants are likely to regulate gene expression. Finally, 96% of the annotated SNPs had minimum chromatin state <8 indicate additional evidence for the regulatory potential of these SNPs.

**UK-Biobank: Chronic Widespread Bodily Pain GWAS cohort of EU ancestry (N=408972)**

**Pain type(s) experienced in last month**

**Fibromyalgia**

**(n=726)**

**ia**

**Neck/Shoulder/**

**Back/Hip/Knee pain**

**for 3 months**

**Headache/Facial/**

**Abdominal pain**

**for 3 months**

**No pain (n=169802)**

**Pain all over the body for 3 months**

**No (n=162590)**

**Yes (n=5440)**

**Yes (n=247722)**

**Reported all 4 pain sites (n=2132)**

**Chronic widespread Bodily pain (n= 6914)** *^a^*

**Controls (n=242929)** *^a^*

**Yes (n=19323)**

**Excluded**

**if reported**

**<4 pain sites**

**Supplementary Figure S1.** Study flowchart of cases and controls.

*^a^*Excluded participants reported to have doctors’ diagnosis of or received nurse diagnosed rheumatoid arthritis (n=4766), polymyalgia rheumatica (n=947), arthritis not otherwise specified (n=3828), systemic lupus erythematosus (n=455), ankylosing spondylitis (n=1187) and myopathy (n = 154).

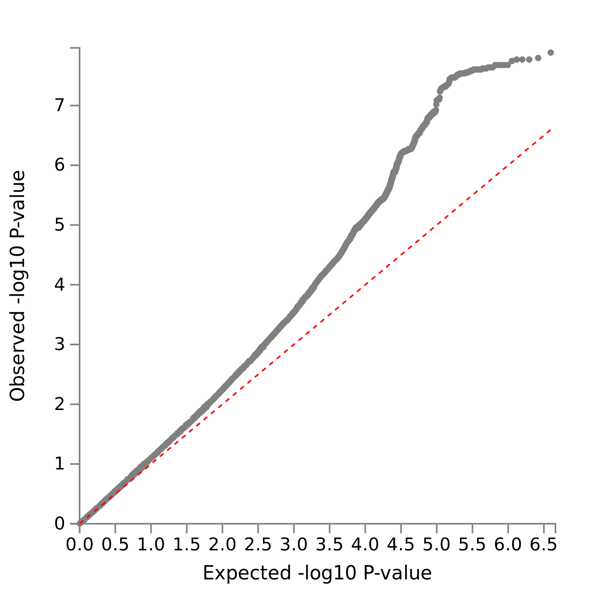

**Supplementary Figure S2.** The QQ plot of GWAS summary statistics of chronic wide-spread pain (CWP) derived from UK biobank European ancestry data. The x-axis displays the expected – log10 transformed p-values and the y-axis displays the observed –log10 transformed p-values.

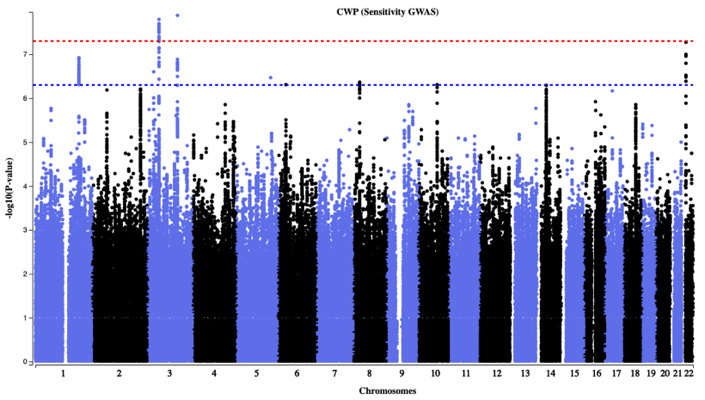

**Supplementary Figure S3.** Manhattan plot of sensitivity GWAS of CWP, which excluded participants who reported chronic non-musculoskeletal pain.

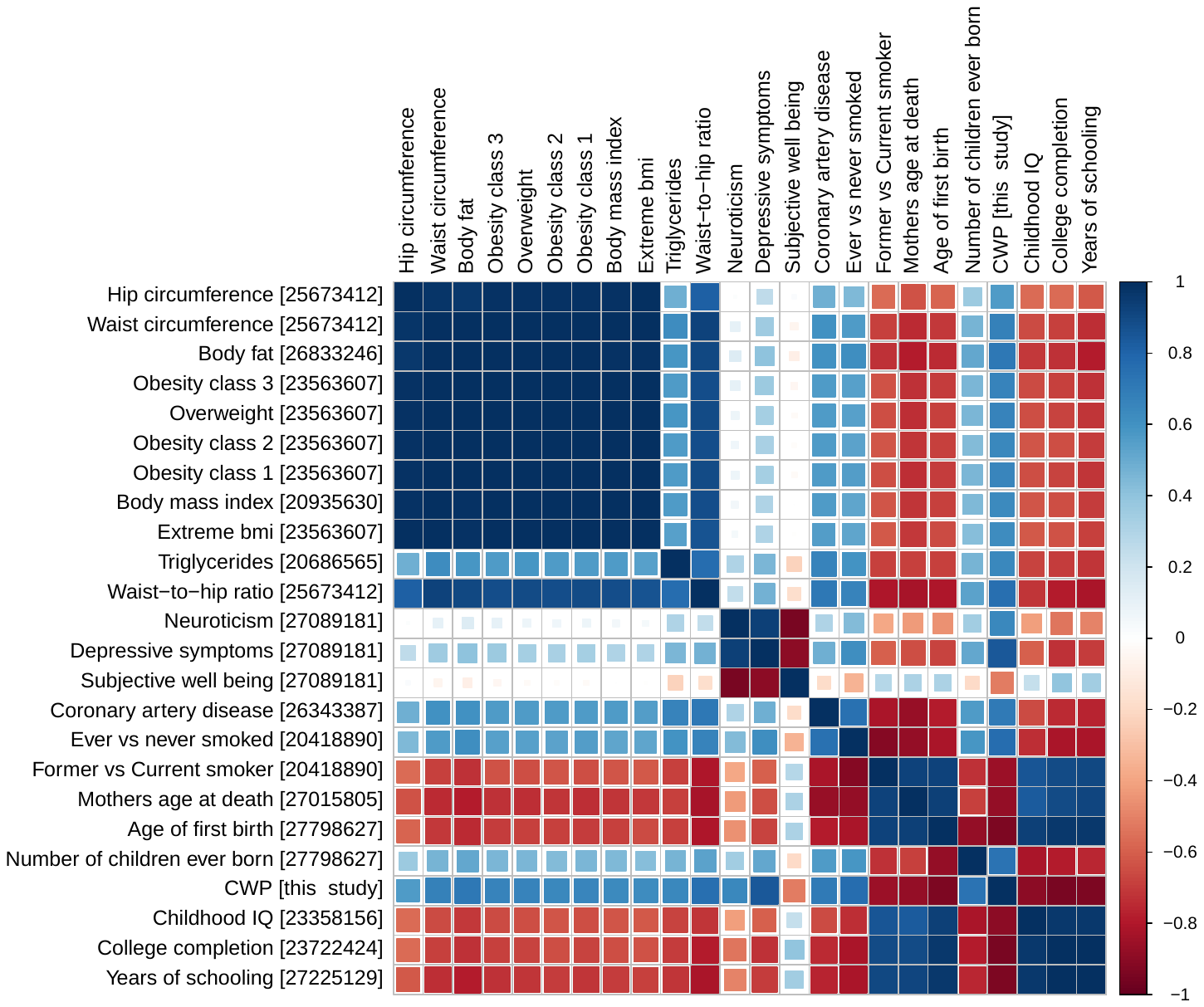

**Supplementary Figure S4.** Heatmap of genetic correlations for 23 complex traits and CWP (absolute rg ≥ 0.20; p < 4.78E-05). Each colored cell indicates magnitudes of genetic correlations. The corresponding color scale is presented on the right side of the heatmap where dark blue represents the highest genetic correlation, and darker red represents highest negative correlation. CWP, chronic widespread pain. On the y-axis, PMID references for each complex trait are placed in the square brackets. On the x-axis, all complex traits are presented maintaining the order of the y-axis.

| A. | B. |
| --- | --- |
| 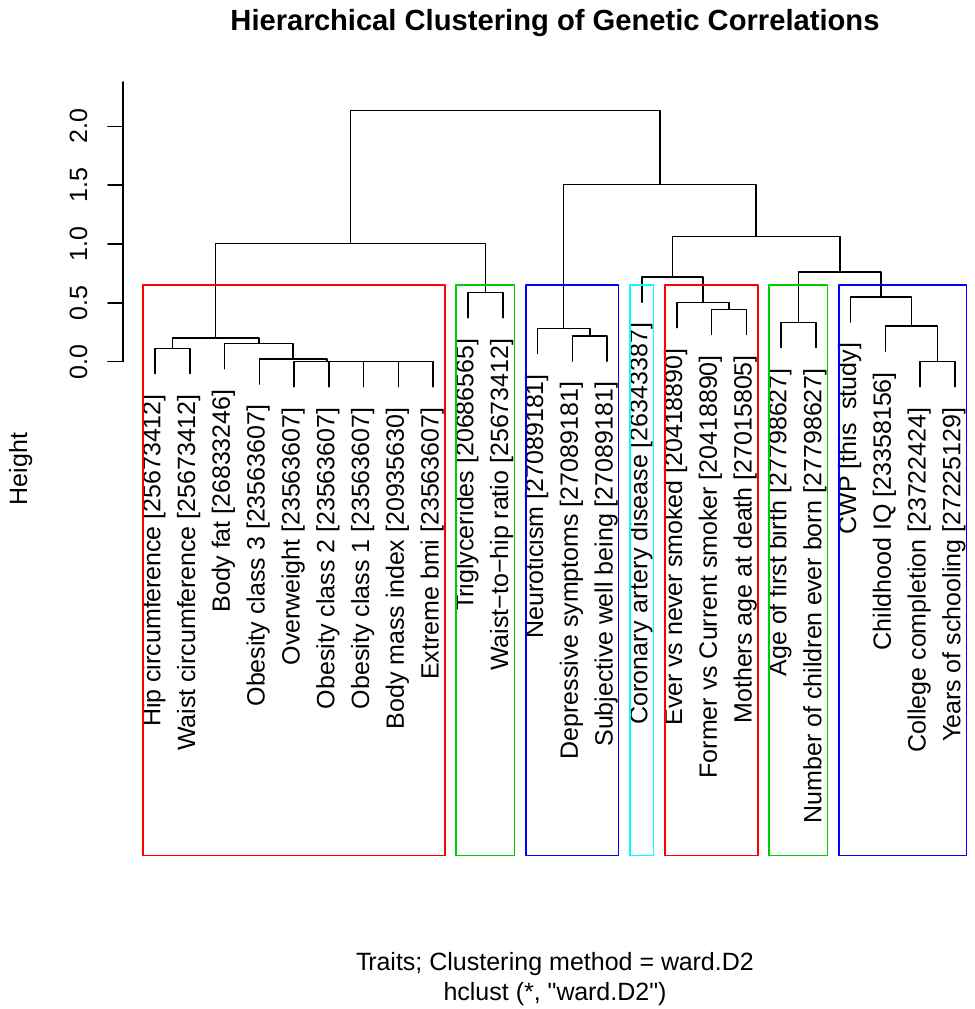 | 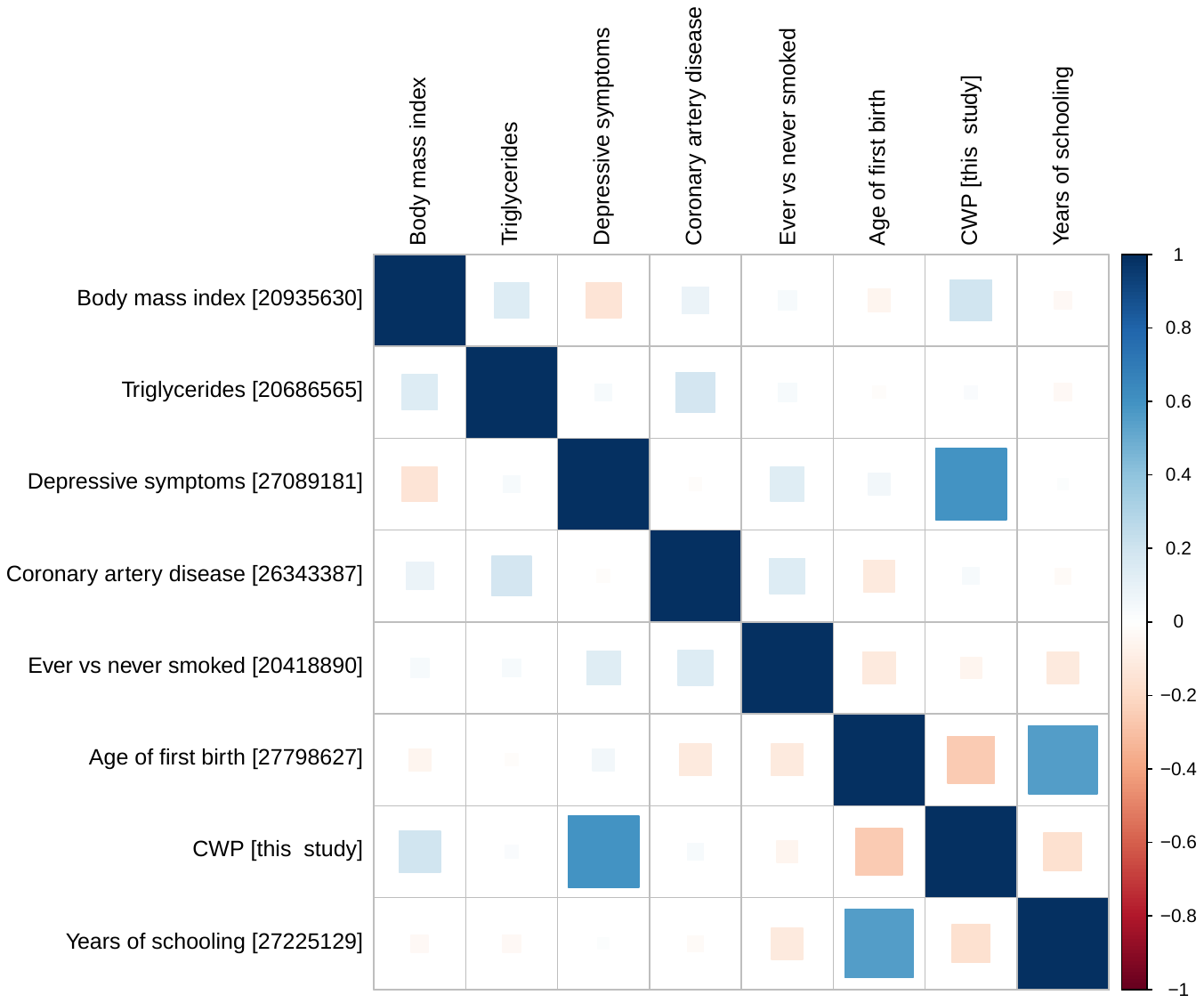 |
| **Supplementary Figure S5.** (A) Hierarchical clustering of genetic correlations for all pairs of traits. PMID references are placed in square brackets. Each cluster indicated with a colored box. A total of 7 clusters were identified and (B) Heatmap of partial genetic correlations for 7 complex traits with CWP. Each colored cell indicates magnitudes of genetic correlations. The corresponding color scale is presented on the right side of the heatmap where dark blue represents the highest genetic correlation, and darker red represents highest negative correlation. CWP, chronic widespread pain. On the y-axis, PMID references for each complex trait are placed in the square brackets. On the x-axis, all complex traits are presented maintaining the order of the y-axis. | |

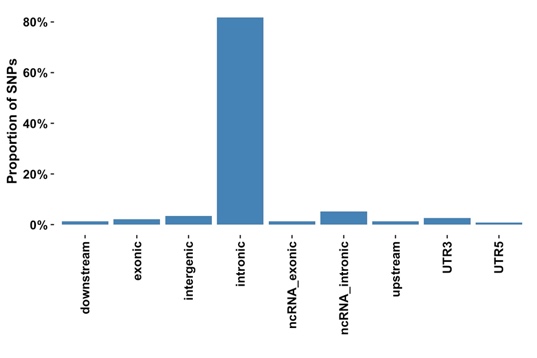

**Supplementary Figure S6.** Functional consequences of candidate SNPs in genomic risk loci annotated by ANNOVAR

| **A.** | **B.** |
| --- | --- |
| 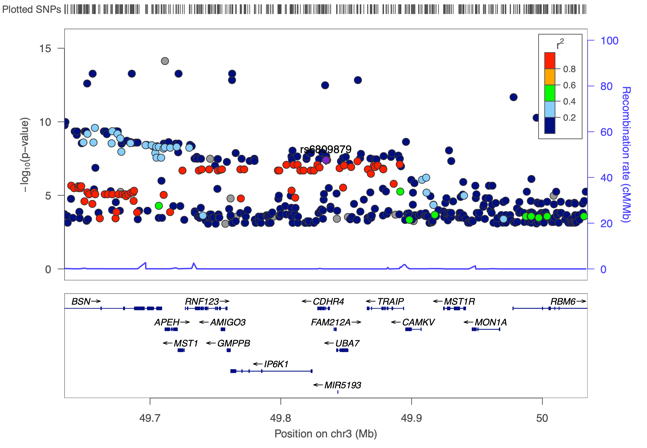 | 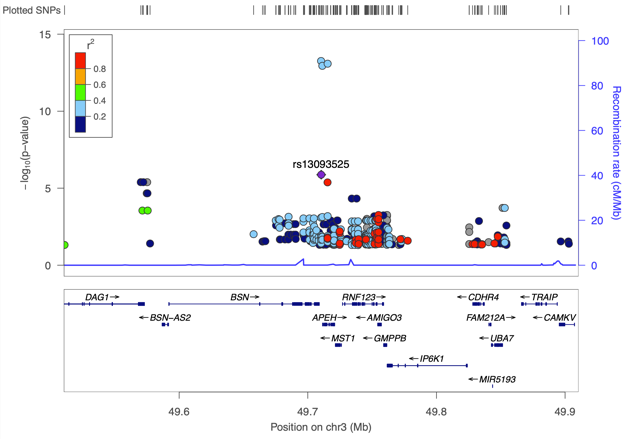 |
| **Supplementary figure S7.** Colocalization of CWP associated locus (*RNF123*) with (A) Skeletal Muscle eQTL (gene-level) and (B) Dorsal root ganglion eQTL (exon-level). Independent SNPs are colored in purple. Other colored circles indicated pairwise LD. Strength of LD (r^2^) presented in the upper right corner of each plot. | |

| **Supplementary Table S1:** Genotyping and imputation methods across all cohorts. | | | |
| --- | --- | --- | --- |
|  | **Genotyping platform** | **Imputation procedure** | **Reference population** |
| **Discovery Cohort** | | | |
| UK Biobank | Applied Biosystems UKB Axiom array, and Applied Biosystems UKB Lung Exome Variant Evaluation Axiom array | IMPUTE 4  MHC: HLA was imputed separately; using HLA*IMP:02 algorithm with a multi-population reference panel. | Haplotype Reference Consortium (HRC), UK10K &1000 genome panel |
| **Replication cohorts** | | | |
| Twins UK | Illumina/HumanHap300, Illumina/HumanHap610Q, Illumina/1M-Duo, and  Illumina/1.2MDuo 1M | MACH | 1000G Phase3 v5 |
| RS-I | Illumina/HumanHap 550K V.3 and Illumina/HumanHap 550K V.3 DUO | MACH | HapMap release 22 CEU |
| RS-II | Illumina/HumanHap 550V.3DUO, and Illumina/HumanHap610Q | MACH | HapMap release 22 CEU |
| RS-III | Illumina/HumanHap610Q | MACH | HapMap release 22 CEU |
| HUNT | Illumina/HumanCoreExome12 v1.0,  Illumina/HumanCoreExome12 v1.1, and UM HUNT Biobank v1.0 | Minimac3 (v2.0.1) | Haplotype Reference Consortium, and HUNT-specific WGS |
| ELSA | Illumina HumanOmni2.5 Bead Chips (HumanOmni2.5-4v1, and HumanOmni2.5-8v1.3) | MACH | Haplotype Reference Consortium |

| **Supplementary Table S2.** Lead SNPs significantly associated with CWP in UK Biobank. | | | | | | | | | | |
| --- | --- | --- | --- | --- | --- | --- | --- | --- | --- | --- |
| **SNP** | **CHR:BP** | **A1** | **A2** | **A1FREQ** | **INFO** | **BETA** | **SE** | **p-value** | **OR, 95% CI** | **Nearest gene** |
| rs1491985 | 3:49739507 | G | C | 0.18 | 1 | 0.0034 | 0.0006 | 1.60E-08 | 1.13, 1.09-1.17 | *RNF123* |
| rs10490825 | 3:130696383 | G | A | 0.87 | 1 | -0.0039 | 0.0007 | 1.30E-08 | 0.87, 0.81-0.93 | *ATP2C1* |
| rs165599 | 22:19956781 | G | A | 0.30 | 1 | -0.0028 | 0.0005 | 2.50E-08 | 0.90, 0.86-0.94 | *COMT/ARVCF* |
| Describe the model and adjustment The independent SNPs at locus reported with RSID, genomic coordinates (CHR:BP; GRCh37.p13/ Hg19/); A1, effect allele; A2, other allele; A1FREQ, effect allele frequency; INFO, estimated imputation score, Beta, linear regression coefficient; SE, standard error; OR, odds ratio; CI, confidence interval, Beta and standard errors of each SNP were divided by (μ * (1 - μ)) to obtain log ORs, where μ represents case fraction. | | | | | | | | | | |

| **Supplementary Table S3.** Results of replication of the three lead SNP by cohort. | | | | | | | | | | | |
| --- | --- | --- | --- | --- | --- | --- | --- | --- | --- | --- | --- |
| **SNP** | **CHR** | **BP** | **A1** | **A2** | **A1FREQ** | **BETA** | **SE** | **p-value** | **N (cases)** | **Power** | **INFO** |
| **TWINS UK** |  |  |  |  |  |  |  |  |  |  |  |
| rs1491985 | 3 | 49739507 | C | G | 0.83 | -0.0348 | 0.0122 | 0.0044 | 4667 (1111) | 5.46% | 1 |
| rs10490825 | 3 | 130696383 | A | G | 0.13 | 0.0091 | 0.0141 | 0.5176 | 4667 (1111) | 5.17% | 0.99 |
| rs165599 | 22 | 19956781 | A | G | 0.69 | 0.0066 | 0.0105 | 0.5290 | 4667 (1111) | 5.30% | 0.99 |
| **HUNT** |  |  |  |  |  |  |  |  |  |  |  |
| rs1491985 | 3 | 49739507 | C | G | 0.8279 | -0.0107 | 0.0062 | 0.0844 | 23795 (10556) | 26.75% | 0.99 |
| rs10490825 | 3 | 130696383 | A | G | 0.1435 | 0.0140 | 0.0067 | 0.0367 | 23795 (10556) | 24.72% | 0.99 |
| rs165599 | 22 | 19956781 | A | G | 0.7196 | 0.0026 | 0.0052 | 0.6162 | 23795 (10556) | 25.68% | Genotyped |
| **ELSA** |  |  |  |  |  |  |  |  |  |  |  |
| rs9870858* | 3 | 49769071 | C | T | 0.1867 | 0.0199 | 0.0092 | 0.0312 | 6983 (1679) | 7.64% | Genotyped |
| rs17329848* | 3 | 130590962 | C | T | 0.1213 | -0.0004 | 0.0110 | 0.9739 | 6983 (1679) | 7.17% | Genotyped |
| rs165599 | 22 | 19956781 | G | A | 0.3049 | -0.0015 | 0.0077 | 0.8433 | 6983 (1679) | 7.39% | Genotyped |
| **RS 1** |  |  |  |  |  |  |  |  |  |  |  |
| rs1491985 | 3 | 49739507 | C | G | 0.8171 | -0.0320 | 0.0899 | 0.7216 | 3136 (532) | 4.12% | 0.98 |
| rs10490825 | 3 | 130696383 | A | G | 0.1296 | 0.1409 | 0.0990 | 0.1549 | 3136 (532) | 3.94% | 1 |
| rs165599 | 22 | 19956781 | A | G | 0.7224 | -0.0042 | 0.0769 | 0.9560 | 3136 (532) | 4.03% | 1 |
| **RS 2** |  |  |  |  |  |  |  |  |  |  |  |
| rs1491985 | 3 | 49739507 | C | G | 0.8077 | -0.01654 | 0.1555 | 0.9153 | 1565 (144) | 2.84% | 1 |
| rs10490825 | 3 | 130696383 | A | G | 0.1383 | 0.005822 | 0.1783 | 0.974 | 1565 (144) | 2.76% | 1 |
| rs165599 | 22 | 19956781 | A | G | 0.7137 | -0.08953 | 0.136 | 0.5103 | 1565 (144) | 2.80% | 1 |
| **RS 3** |  |  |  |  |  |  |  |  |  |  |  |
| rs1491985 | 3 | 49739507 | C | G | 0.8047 | -0.2453 | 0.1381 | 0.0758 | 2934 (155) | 3.95% | 1 |
| rs10490825 | 3 | 130696383 | A | G | 0.1377 | 0.0877 | 0.1685 | 0.6027 | 2934 (155) | 3.78% | 0.99 |
| rs165599 | 22 | 19956781 | A | G | 0.7217 | -0.0905 | 0.1303 | 0.4874 | 2934 (155) | 3.86% | 0.99 |
| Replication SNPs at locus reported with RSID, genomic coordinates (CHR:BP; GRCh37.p13/ Hg19/); A1, effect allele; A2, Alternative allele; A1FREQ, effect allele frequency; Beta, linear regression coefficient; SE, standard error; N, sample size used of each SNP analysis; INFO, Imputation score; HUNT: The Nord-Trøndelag Health Study; ELSA: The English Longitudinal Study of Ageing; RS-1, 2 and 3: The Rotterdam Study 1,2,3.  *rs9870858 (instead of rs1491985) and rs17329848 (instead of rs10490825) were used as proxy in the ELSA cohort. | | | | | | | | | | | |

| **Supplementary Table S4.** Sample-size based meta-analysis findings of replication SNPs. | | | | | | | | | | | | |
| --- | --- | --- | --- | --- | --- | --- | --- | --- | --- | --- | --- | --- |
| **SNP** | **A1** | **A2** | **A1Freq** | **FreqSE** | **Weight** | **Z** | **P-value** | **Direction** | **HetI^2^** | **HetChi^2^** | **HetPVal** | **Power** |
| rs1491985* | C | G | 0.82 | 0.008 | 43080 | -3.667 | 0.0002 | ------ | 10.20 | 5.57 | 0.35 | 49.70 |
| rs10490825* | A | G | 0.14 | 0.0083 | 43080 | 2.278 | 0.0227 | ++-+++ | 0 | 1.89 | 0.86 | 46.32 |
| rs165599 | A | G | 0.71 | 0.0124 | 43080 | 0.338 | 0.7356 | +++--- | 0 | 1.49 | 0.91 | 47.93 |
| A1Freq: Frequency of A1 allele; FreqSE: Standard error for the frequency of A1 allele; Z: Z statistics; HetI^2^: Heterogeneity I2 parameter; HetChi^2^: Heterogeneity test statistic; HetPVal: P-value for heterogeneity statistic.  * rs9870858 (instead of rs1491985) and rs17329848 (instead of rs10490825) were used as proxy in the ELSA cohort. | | | | | | | | | | | | |

| **Supplementary Table S5.** Standard error based meta-analysis findings of replication SNPs. | | | | | | | | | | | |
| --- | --- | --- | --- | --- | --- | --- | --- | --- | --- | --- | --- |
| **SNP** | **A1** | **A2** | **A1Freq** | **FreqSE** | **Effect** | **StdErr** | **P-value** | **Direction** | **HetI^2^** | **HetChi^2^** | **HetPVal** |
| rs1491985* | C | G | 0.82 | 0.0064 | -0.0171 | 0.0047 | 0.0003 | ------ | 17 | 6.03 | 0.30 |
| rs10490825* | A | G | 0.14 | 0.0093 | 0.0104 | 0.0053 | 0.049 | ++-+++ | 0 | 3.21 | 0.67 |
| rs165599 | A | G | 0.71 | 0.0134 | 0.0027 | 0.004 | 0.50 | +++--- | 0 | 1.14 | 0.95 |
| A1Freq: Frequency of A1 allele; FreqSE: Standard error for the frequency of A1 allele; StdErr: Standard Error, HetI^2^: Heterogeneity I2 parameter; HetChi^2^: Heterogeneity test statistic; HetPVal: P-value for heterogeneity statistic.  * rs9870858 (instead of rs1491985) and rs17329848 (instead of rs10490825) were used as proxy in the ELSA cohort. | | | | | | | | | | | |

| **Supplementary Table S6.** Genetic and partial genetic correlations for chronic widespread musculoskeletal pain with body mass index, triglycerides, depressive symptoms, coronary artery disease, ever vs never smoked, age of first birth and years of schooling. | | | | | |
| --- | --- | --- | --- | --- | --- |
|  | Genetic correlations | | | Partial genetic correlations | |
|  | rg | SE | P | Partial rg | P |
| Body mass index [20935630] | 0.31 | 0.0358 | 8.81E-18 | 0.2 | 2.36E-08 |
| Triglycerides [20686565] | 0.20 | 0.0462 | 1.10E-05 | 0.02 | 0.6571 |
| Depressive symptoms [27089181] | 0.65 | 0.0516 | 2.06E-36 | 0.59 | 9.22E-31 |
| Coronary artery disease [26343387] | 0.25 | 0.0397 | 5.38E-10 | 0.03 | 0.4293 |
| Ever vs never smoked [20418890] | 0.27 | 0.0588 | 5.83E-06 | -0.05 | 0.3802 |
| Age of first birth [27798627] | -0.58 | 0.0412 | 2.03E-44 | -0.26 | 3.24E-10 |
| Years of schooling [27225129] | -0.54 | 0.0331 | 4.23E-60 | -0.17 | 3.37E-07 |
| rg, genetic correlation estimate; SE, standard error; P, p-value; partial rg, partial genetic correlation estimate. | | | | | |

| **Supplementary table S7.** Differential gene set enrichment in 54 specific tissue types from GTEx. | | | | | | |
| --- | --- | --- | --- | --- | --- | --- |
| Category | GeneSet | N_genes | N_overlap | p | adjP | genes |
| DEG.twoside | Muscle_Skeletal | 7979 | 51 | 3.27E-10 | 1.76E-08 | ENSG00000115365:ENSG00000100075:ENSG00000070371:ENSG00000184058:ENSG00000215012:ENSG00000093010:ENSG00000099889:ENSG00000128191:ENSG00000099899:ENSG00000099901:ENSG00000099904:ENSG00000213672:ENSG00000114302:ENSG00000178537:ENSG00000177479:ENSG00000178467:ENSG00000178149:ENSG00000198218:ENSG00000177352:ENSG00000188315:ENSG00000114316:ENSG00000145022:ENSG00000145020:ENSG00000145029:ENSG00000173531:ENSG00000164068:ENSG00000173540:ENSG00000176095:ENSG00000185614:ENSG00000182179:ENSG00000183763:ENSG00000004534:ENSG00000001617:ENSG00000114353:ENSG00000243477:ENSG00000114378:ENSG00000068001:ENSG00000007402:ENSG00000114735:ENSG00000114738:ENSG00000088538:ENSG00000196455:ENSG00000017260:ENSG00000034533:ENSG00000198585:ENSG00000138246:ENSG00000240303:ENSG00000113971:ENSG00000125388:ENSG00000004864:ENSG00000092964 |
| DEG.down | Brain_Hippocampus | 7493 | 48 | 1.78E-09 | 9.63E-08 | ENSG00000021826:ENSG00000070371:ENSG00000070010:ENSG00000215012:ENSG00000099889:ENSG00000183597:ENSG00000128191:ENSG00000099899:ENSG00000099901:ENSG00000099904:ENSG00000234409:ENSG00000099917:ENSG00000114302:ENSG00000178537:ENSG00000177479:ENSG00000178252:ENSG00000178149:ENSG00000198218:ENSG00000172037:ENSG00000177352:ENSG00000185909:ENSG00000188315:ENSG00000114316:ENSG00000145020:ENSG00000173402:ENSG00000164062:ENSG00000173531:ENSG00000164068:ENSG00000173540:ENSG00000185614:ENSG00000182179:ENSG00000183763:ENSG00000001617:ENSG00000214706:ENSG00000243477:ENSG00000114378:ENSG00000068001:ENSG00000007402:ENSG00000114735:ENSG00000114738:ENSG00000196455:ENSG00000034533:ENSG00000114686:ENSG00000138246:ENSG00000240303:ENSG00000113971:ENSG00000124664:ENSG00000004864 |
| DEG.down | Whole_Blood | 6908 | 45 | 5.47E-09 | 2.95E-07 | ENSG00000115365:ENSG00000100075:ENSG00000070371:ENSG00000100084:ENSG00000215012:ENSG00000093010:ENSG00000099889:ENSG00000128191:ENSG00000099899:ENSG00000099901:ENSG00000099904:ENSG00000213672:ENSG00000114302:ENSG00000177479:ENSG00000178467:ENSG00000178252:ENSG00000178149:ENSG00000198218:ENSG00000172037:ENSG00000177352:ENSG00000185909:ENSG00000145020:ENSG00000145029:ENSG00000173402:ENSG00000164062:ENSG00000173531:ENSG00000173540:ENSG00000185614:ENSG00000164077:ENSG00000004534:ENSG00000214706:ENSG00000243477:ENSG00000068001:ENSG00000114383:ENSG00000007402:ENSG00000114735:ENSG00000196455:ENSG00000017260:ENSG00000034533:ENSG00000114686:ENSG00000138246:ENSG00000240303:ENSG00000113971:ENSG00000004864:ENSG00000092964 |
| DEG.down | Muscle_Skeletal | 6836 | 44 | 1.39E-08 | 7.50E-07 | ENSG00000115365:ENSG00000100075:ENSG00000215012:ENSG00000093010:ENSG00000099889:ENSG00000128191:ENSG00000099899:ENSG00000099901:ENSG00000099904:ENSG00000213672:ENSG00000178467:ENSG00000178149:ENSG00000198218:ENSG00000177352:ENSG00000188315:ENSG00000114316:ENSG00000145022:ENSG00000145020:ENSG00000145029:ENSG00000173531:ENSG00000173540:ENSG00000176095:ENSG00000185614:ENSG00000182179:ENSG00000183763:ENSG00000004534:ENSG00000001617:ENSG00000114353:ENSG00000243477:ENSG00000114378:ENSG00000068001:ENSG00000007402:ENSG00000114735:ENSG00000088538:ENSG00000196455:ENSG00000017260:ENSG00000034533:ENSG00000198585:ENSG00000138246:ENSG00000240303:ENSG00000113971:ENSG00000125388:ENSG00000004864:ENSG00000092964 |
| DEG.down | Brain_Amygdala | 7678 | 46 | 5.01E-08 | 2.70E-06 | ENSG00000021826:ENSG00000070371:ENSG00000070010:ENSG00000215012:ENSG00000183597:ENSG00000128191:ENSG00000099899:ENSG00000099904:ENSG00000099917:ENSG00000114302:ENSG00000178537:ENSG00000177479:ENSG00000178252:ENSG00000178149:ENSG00000198218:ENSG00000172037:ENSG00000177352:ENSG00000185909:ENSG00000188315:ENSG00000114316:ENSG00000145020:ENSG00000173402:ENSG00000164062:ENSG00000173531:ENSG00000164068:ENSG00000173540:ENSG00000185614:ENSG00000182179:ENSG00000183763:ENSG00000004534:ENSG00000003756:ENSG00000001617:ENSG00000214706:ENSG00000243477:ENSG00000114378:ENSG00000068001:ENSG00000007402:ENSG00000114735:ENSG00000114738:ENSG00000196455:ENSG00000034533:ENSG00000114686:ENSG00000138246:ENSG00000240303:ENSG00000113971:ENSG00000004864 |
| DEG.down | Brain_Putamen_basal_ganglia | 7776 | 46 | 7.54E-08 | 4.07E-06 | ENSG00000070371:ENSG00000070010:ENSG00000215012:ENSG00000099889:ENSG00000183597:ENSG00000128191:ENSG00000099899:ENSG00000099901:ENSG00000099904:ENSG00000234409:ENSG00000099917:ENSG00000114302:ENSG00000178537:ENSG00000177479:ENSG00000178252:ENSG00000178149:ENSG00000198218:ENSG00000172037:ENSG00000177352:ENSG00000185909:ENSG00000188315:ENSG00000114316:ENSG00000145020:ENSG00000145029:ENSG00000173402:ENSG00000164062:ENSG00000173531:ENSG00000164068:ENSG00000173540:ENSG00000185614:ENSG00000182179:ENSG00000004534:ENSG00000003756:ENSG00000001617:ENSG00000214706:ENSG00000114378:ENSG00000068001:ENSG00000114735:ENSG00000114738:ENSG00000196455:ENSG00000034533:ENSG00000114686:ENSG00000138246:ENSG00000240303:ENSG00000113971:ENSG00000004864 |
| DEG.twoside | Brain_Hippocampus | 9458 | 51 | 1.69E-07 | 9.11E-06 | ENSG00000021826:ENSG00000070371:ENSG00000070010:ENSG00000215012:ENSG00000099889:ENSG00000183597:ENSG00000128191:ENSG00000099899:ENSG00000099901:ENSG00000099904:ENSG00000234409:ENSG00000099917:ENSG00000114302:ENSG00000178537:ENSG00000177479:ENSG00000178252:ENSG00000178149:ENSG00000198218:ENSG00000172037:ENSG00000177352:ENSG00000185909:ENSG00000188315:ENSG00000114316:ENSG00000145020:ENSG00000173402:ENSG00000164061:ENSG00000164062:ENSG00000173531:ENSG00000164068:ENSG00000173540:ENSG00000185614:ENSG00000182179:ENSG00000183763:ENSG00000164076:ENSG00000001617:ENSG00000214706:ENSG00000243477:ENSG00000114378:ENSG00000068001:ENSG00000007402:ENSG00000114735:ENSG00000114738:ENSG00000088538:ENSG00000196455:ENSG00000034533:ENSG00000114686:ENSG00000138246:ENSG00000240303:ENSG00000113971:ENSG00000124664:ENSG00000004864 |
| DEG.down | Brain_Nucleus_accumbens_basal_ganglia | 6409 | 39 | 7.99E-07 | 4.32E-05 | ENSG00000070371:ENSG00000070010:ENSG00000215012:ENSG00000099889:ENSG00000183597:ENSG00000128191:ENSG00000099899:ENSG00000099901:ENSG00000099904:ENSG00000099917:ENSG00000114302:ENSG00000178537:ENSG00000177479:ENSG00000178149:ENSG00000198218:ENSG00000172037:ENSG00000177352:ENSG00000185909:ENSG00000188315:ENSG00000114316:ENSG00000145020:ENSG00000173402:ENSG00000173531:ENSG00000164068:ENSG00000173540:ENSG00000185614:ENSG00000182179:ENSG00000001617:ENSG00000214706:ENSG00000114378:ENSG00000068001:ENSG00000114735:ENSG00000114738:ENSG00000196455:ENSG00000034533:ENSG00000138246:ENSG00000240303:ENSG00000113971:ENSG00000004864 |
| DEG.twoside | Heart_Left_Ventricle | 9979 | 51 | 1.06E-06 | 5.74E-05 | ENSG00000115365:ENSG00000100075:ENSG00000070371:ENSG00000100084:ENSG00000070010:ENSG00000215012:ENSG00000093010:ENSG00000099889:ENSG00000128191:ENSG00000099899:ENSG00000099901:ENSG00000099904:ENSG00000099917:ENSG00000213672:ENSG00000114302:ENSG00000178467:ENSG00000178252:ENSG00000178149:ENSG00000198218:ENSG00000177352:ENSG00000188315:ENSG00000114316:ENSG00000145022:ENSG00000145020:ENSG00000145029:ENSG00000173531:ENSG00000164068:ENSG00000173540:ENSG00000176095:ENSG00000185614:ENSG00000182179:ENSG00000164077:ENSG00000003756:ENSG00000001617:ENSG00000243477:ENSG00000114378:ENSG00000114383:ENSG00000114735:ENSG00000114738:ENSG00000088538:ENSG00000196455:ENSG00000017260:ENSG00000034533:ENSG00000114686:ENSG00000138246:ENSG00000240303:ENSG00000113971:ENSG00000129048:ENSG00000125388:ENSG00000004864:ENSG00000092964 |
| DEG.down | Heart_Left_Ventricle | 9443 | 49 | 1.41E-06 | 7.61E-05 | ENSG00000115365:ENSG00000100075:ENSG00000070371:ENSG00000100084:ENSG00000070010:ENSG00000215012:ENSG00000093010:ENSG00000099889:ENSG00000128191:ENSG00000099899:ENSG00000099901:ENSG00000099904:ENSG00000099917:ENSG00000213672:ENSG00000114302:ENSG00000178467:ENSG00000178252:ENSG00000178149:ENSG00000198218:ENSG00000177352:ENSG00000188315:ENSG00000114316:ENSG00000145022:ENSG00000145020:ENSG00000145029:ENSG00000173531:ENSG00000164068:ENSG00000173540:ENSG00000176095:ENSG00000185614:ENSG00000182179:ENSG00000164077:ENSG00000003756:ENSG00000001617:ENSG00000243477:ENSG00000114383:ENSG00000114735:ENSG00000088538:ENSG00000196455:ENSG00000017260:ENSG00000034533:ENSG00000114686:ENSG00000138246:ENSG00000240303:ENSG00000113971:ENSG00000129048:ENSG00000125388:ENSG00000004864:ENSG00000092964 |
| DEG.twoside | Brain_Nucleus_accumbens_basal_ganglia | 8860 | 47 | 1.53E-06 | 8.26E-05 | ENSG00000070371:ENSG00000070010:ENSG00000215012:ENSG00000099889:ENSG00000183597:ENSG00000128191:ENSG00000099899:ENSG00000099901:ENSG00000099904:ENSG00000099917:ENSG00000008300:ENSG00000114302:ENSG00000178537:ENSG00000177479:ENSG00000178467:ENSG00000178149:ENSG00000198218:ENSG00000172037:ENSG00000177352:ENSG00000185909:ENSG00000188315:ENSG00000114316:ENSG00000145020:ENSG00000173402:ENSG00000164061:ENSG00000173531:ENSG00000164068:ENSG00000173540:ENSG00000185614:ENSG00000182179:ENSG00000183763:ENSG00000164076:ENSG00000001617:ENSG00000214706:ENSG00000114378:ENSG00000068001:ENSG00000007402:ENSG00000114735:ENSG00000114738:ENSG00000088538:ENSG00000196455:ENSG00000034533:ENSG00000114670:ENSG00000138246:ENSG00000240303:ENSG00000113971:ENSG00000004864 |
| DEG.down | Brain_Substantia_nigra | 7162 | 41 | 1.79E-06 | 9.66E-05 | ENSG00000070371:ENSG00000070010:ENSG00000215012:ENSG00000099889:ENSG00000128191:ENSG00000099899:ENSG00000099904:ENSG00000234409:ENSG00000099917:ENSG00000114302:ENSG00000178537:ENSG00000177479:ENSG00000178252:ENSG00000178149:ENSG00000198218:ENSG00000172037:ENSG00000177352:ENSG00000185909:ENSG00000188315:ENSG00000114316:ENSG00000145020:ENSG00000145029:ENSG00000173402:ENSG00000164062:ENSG00000173531:ENSG00000164068:ENSG00000173540:ENSG00000176095:ENSG00000185614:ENSG00000182179:ENSG00000001617:ENSG00000214706:ENSG00000243477:ENSG00000114378:ENSG00000068001:ENSG00000114735:ENSG00000114738:ENSG00000196455:ENSG00000034533:ENSG00000138246:ENSG00000113971 |
| DEG.twoside | Whole_Blood | 8941 | 47 | 2.02E-06 | 1.09E-04 | ENSG00000115365:ENSG00000100075:ENSG00000070371:ENSG00000100084:ENSG00000215012:ENSG00000093010:ENSG00000099889:ENSG00000183597:ENSG00000128191:ENSG00000099899:ENSG00000099901:ENSG00000099904:ENSG00000213672:ENSG00000114302:ENSG00000177479:ENSG00000178467:ENSG00000178252:ENSG00000178149:ENSG00000198218:ENSG00000172037:ENSG00000177352:ENSG00000185909:ENSG00000188315:ENSG00000145020:ENSG00000145029:ENSG00000173402:ENSG00000164062:ENSG00000173531:ENSG00000173540:ENSG00000185614:ENSG00000164077:ENSG00000004534:ENSG00000214706:ENSG00000243477:ENSG00000068001:ENSG00000114383:ENSG00000007402:ENSG00000114735:ENSG00000196455:ENSG00000017260:ENSG00000034533:ENSG00000114686:ENSG00000138246:ENSG00000240303:ENSG00000113971:ENSG00000004864:ENSG00000092964 |
| DEG.down | Brain_Spinal_cord_cervical_c-1 | 5083 | 33 | 2.26E-06 | 1.22E-04 | ENSG00000070371:ENSG00000215012:ENSG00000099889:ENSG00000128191:ENSG00000099904:ENSG00000234409:ENSG00000099917:ENSG00000114302:ENSG00000178149:ENSG00000198218:ENSG00000172037:ENSG00000177352:ENSG00000185909:ENSG00000114316:ENSG00000145020:ENSG00000145029:ENSG00000173402:ENSG00000164062:ENSG00000173531:ENSG00000164068:ENSG00000173540:ENSG00000185614:ENSG00000182179:ENSG00000183763:ENSG00000001617:ENSG00000214706:ENSG00000186792:ENSG00000114378:ENSG00000068001:ENSG00000114735:ENSG00000114738:ENSG00000196455:ENSG00000138246 |
| DEG.down | Pancreas | 9586 | 49 | 2.28E-06 | 1.23E-04 | ENSG00000115365:ENSG00000070371:ENSG00000100084:ENSG00000070010:ENSG00000215012:ENSG00000093010:ENSG00000099889:ENSG00000183597:ENSG00000128191:ENSG00000099899:ENSG00000099901:ENSG00000099904:ENSG00000099917:ENSG00000213672:ENSG00000114302:ENSG00000178537:ENSG00000177479:ENSG00000178057:ENSG00000198218:ENSG00000177352:ENSG00000185909:ENSG00000188315:ENSG00000114316:ENSG00000145022:ENSG00000145029:ENSG00000173531:ENSG00000164068:ENSG00000176095:ENSG00000182179:ENSG00000164077:ENSG00000001617:ENSG00000114353:ENSG00000186792:ENSG00000243477:ENSG00000114378:ENSG00000068001:ENSG00000114383:ENSG00000007402:ENSG00000196455:ENSG00000017260:ENSG00000034533:ENSG00000114670:ENSG00000198585:ENSG00000138246:ENSG00000240303:ENSG00000113971:ENSG00000125388:ENSG00000004864:ENSG00000092964 |
| DEG.down | Brain_Caudate_basal_ganglia | 6858 | 39 | 4.66E-06 | 2.51E-04 | ENSG00000070371:ENSG00000070010:ENSG00000215012:ENSG00000099889:ENSG00000183597:ENSG00000128191:ENSG00000099899:ENSG00000099901:ENSG00000099904:ENSG00000234409:ENSG00000099917:ENSG00000114302:ENSG00000178537:ENSG00000177479:ENSG00000178149:ENSG00000198218:ENSG00000172037:ENSG00000177352:ENSG00000185909:ENSG00000145020:ENSG00000173402:ENSG00000164062:ENSG00000173531:ENSG00000164068:ENSG00000173540:ENSG00000185614:ENSG00000182179:ENSG00000001617:ENSG00000214706:ENSG00000114378:ENSG00000068001:ENSG00000114735:ENSG00000114738:ENSG00000196455:ENSG00000034533:ENSG00000138246:ENSG00000240303:ENSG00000113971:ENSG00000004864 |
| DEG.twoside | Brain_Amygdala | 9498 | 48 | 4.69E-06 | 2.53E-04 | ENSG00000021826:ENSG00000070371:ENSG00000070010:ENSG00000215012:ENSG00000183597:ENSG00000128191:ENSG00000099899:ENSG00000099904:ENSG00000099917:ENSG00000114302:ENSG00000178537:ENSG00000177479:ENSG00000178252:ENSG00000178149:ENSG00000198218:ENSG00000172037:ENSG00000177352:ENSG00000185909:ENSG00000188315:ENSG00000114316:ENSG00000145020:ENSG00000173402:ENSG00000164061:ENSG00000164062:ENSG00000173531:ENSG00000164068:ENSG00000173540:ENSG00000185614:ENSG00000182179:ENSG00000183763:ENSG00000164076:ENSG00000004534:ENSG00000003756:ENSG00000001617:ENSG00000214706:ENSG00000243477:ENSG00000114378:ENSG00000068001:ENSG00000007402:ENSG00000114735:ENSG00000114738:ENSG00000196455:ENSG00000034533:ENSG00000114686:ENSG00000138246:ENSG00000240303:ENSG00000113971:ENSG00000004864 |
| DEG.twoside | Brain_Putamen_basal_ganglia | 9517 | 48 | 4.99E-06 | 2.69E-04 | ENSG00000070371:ENSG00000070010:ENSG00000215012:ENSG00000099889:ENSG00000183597:ENSG00000128191:ENSG00000099899:ENSG00000099901:ENSG00000099904:ENSG00000234409:ENSG00000099917:ENSG00000114302:ENSG00000178537:ENSG00000177479:ENSG00000178252:ENSG00000178149:ENSG00000198218:ENSG00000172037:ENSG00000177352:ENSG00000185909:ENSG00000188315:ENSG00000114316:ENSG00000145020:ENSG00000145029:ENSG00000173402:ENSG00000164061:ENSG00000164062:ENSG00000173531:ENSG00000164068:ENSG00000173540:ENSG00000185614:ENSG00000182179:ENSG00000164076:ENSG00000004534:ENSG00000003756:ENSG00000001617:ENSG00000214706:ENSG00000114378:ENSG00000068001:ENSG00000114735:ENSG00000114738:ENSG00000196455:ENSG00000034533:ENSG00000114686:ENSG00000138246:ENSG00000240303:ENSG00000113971:ENSG00000004864 |
| DEG.down | Brain_Anterior_cingulate_cortex_BA24 | 6651 | 38 | 6.05E-06 | 3.27E-04 | ENSG00000021826:ENSG00000070371:ENSG00000070010:ENSG00000128191:ENSG00000099899:ENSG00000234409:ENSG00000099917:ENSG00000114302:ENSG00000178537:ENSG00000177479:ENSG00000178149:ENSG00000198218:ENSG00000172037:ENSG00000177352:ENSG00000185909:ENSG00000188315:ENSG00000114316:ENSG00000145020:ENSG00000173402:ENSG00000173531:ENSG00000164068:ENSG00000173540:ENSG00000185614:ENSG00000182179:ENSG00000183763:ENSG00000001617:ENSG00000214706:ENSG00000114378:ENSG00000068001:ENSG00000114735:ENSG00000114738:ENSG00000196455:ENSG00000034533:ENSG00000138246:ENSG00000240303:ENSG00000113971:ENSG00000124664:ENSG00000004864 |
| DEG.twoside | Pancreas | 10331 | 49 | 2.30E-05 | 1.24E-03 | ENSG00000115365:ENSG00000070371:ENSG00000100084:ENSG00000070010:ENSG00000215012:ENSG00000093010:ENSG00000099889:ENSG00000183597:ENSG00000128191:ENSG00000099899:ENSG00000099901:ENSG00000099904:ENSG00000099917:ENSG00000213672:ENSG00000114302:ENSG00000178537:ENSG00000177479:ENSG00000178057:ENSG00000198218:ENSG00000177352:ENSG00000185909:ENSG00000188315:ENSG00000114316:ENSG00000145022:ENSG00000145029:ENSG00000173531:ENSG00000164068:ENSG00000176095:ENSG00000182179:ENSG00000164077:ENSG00000001617:ENSG00000114353:ENSG00000186792:ENSG00000243477:ENSG00000114378:ENSG00000068001:ENSG00000114383:ENSG00000007402:ENSG00000196455:ENSG00000017260:ENSG00000034533:ENSG00000114670:ENSG00000198585:ENSG00000138246:ENSG00000240303:ENSG00000113971:ENSG00000125388:ENSG00000004864:ENSG00000092964 |
| DEG.down | Heart_Atrial_Appendage | 8409 | 42 | 4.59E-05 | 2.48E-03 | ENSG00000115365:ENSG00000021826:ENSG00000100075:ENSG00000100084:ENSG00000070010:ENSG00000215012:ENSG00000093010:ENSG00000099889:ENSG00000128191:ENSG00000099899:ENSG00000099901:ENSG00000099904:ENSG00000099917:ENSG00000114302:ENSG00000178467:ENSG00000178149:ENSG00000198218:ENSG00000177352:ENSG00000188315:ENSG00000114316:ENSG00000145022:ENSG00000145029:ENSG00000173531:ENSG00000164068:ENSG00000173540:ENSG00000176095:ENSG00000185614:ENSG00000182179:ENSG00000164077:ENSG00000001617:ENSG00000186792:ENSG00000243477:ENSG00000114735:ENSG00000088538:ENSG00000196455:ENSG00000017260:ENSG00000034533:ENSG00000114670:ENSG00000198585:ENSG00000138246:ENSG00000113971:ENSG00000004864 |
| DEG.twoside | Brain_Caudate_basal_ganglia | 9070 | 44 | 5.41E-05 | 2.92E-03 | ENSG00000070371:ENSG00000070010:ENSG00000215012:ENSG00000099889:ENSG00000183597:ENSG00000128191:ENSG00000099899:ENSG00000099901:ENSG00000099904:ENSG00000234409:ENSG00000099917:ENSG00000114302:ENSG00000178537:ENSG00000177479:ENSG00000178149:ENSG00000198218:ENSG00000172037:ENSG00000177352:ENSG00000185909:ENSG00000145020:ENSG00000173402:ENSG00000164061:ENSG00000164062:ENSG00000173531:ENSG00000164068:ENSG00000173540:ENSG00000185614:ENSG00000182179:ENSG00000164076:ENSG00000001617:ENSG00000214706:ENSG00000114378:ENSG00000068001:ENSG00000007402:ENSG00000114735:ENSG00000114738:ENSG00000088538:ENSG00000196455:ENSG00000034533:ENSG00000114670:ENSG00000138246:ENSG00000240303:ENSG00000113971:ENSG00000004864 |
| DEG.twoside | Brain_Spinal_cord_cervical_c-1 | 7307 | 38 | 5.74E-05 | 3.10E-03 | ENSG00000115365:ENSG00000021826:ENSG00000070371:ENSG00000215012:ENSG00000099889:ENSG00000128191:ENSG00000099904:ENSG00000234409:ENSG00000099917:ENSG00000114302:ENSG00000178149:ENSG00000198218:ENSG00000172037:ENSG00000177352:ENSG00000185909:ENSG00000114316:ENSG00000145020:ENSG00000145029:ENSG00000173402:ENSG00000164062:ENSG00000173531:ENSG00000164068:ENSG00000173540:ENSG00000185614:ENSG00000182179:ENSG00000183763:ENSG00000001617:ENSG00000214706:ENSG00000186792:ENSG00000114378:ENSG00000068001:ENSG00000114735:ENSG00000114738:ENSG00000088538:ENSG00000196455:ENSG00000198585:ENSG00000138246:ENSG00000004864 |
| DEG.twoside | Brain_Substantia_nigra | 8807 | 43 | 6.11E-05 | 3.30E-03 | ENSG00000021826:ENSG00000070371:ENSG00000070010:ENSG00000215012:ENSG00000099889:ENSG00000128191:ENSG00000099899:ENSG00000099904:ENSG00000234409:ENSG00000099917:ENSG00000114302:ENSG00000178537:ENSG00000177479:ENSG00000178252:ENSG00000178149:ENSG00000198218:ENSG00000172037:ENSG00000177352:ENSG00000185909:ENSG00000188315:ENSG00000114316:ENSG00000145020:ENSG00000145029:ENSG00000173402:ENSG00000164062:ENSG00000173531:ENSG00000164068:ENSG00000173540:ENSG00000176095:ENSG00000185614:ENSG00000182179:ENSG00000001617:ENSG00000214706:ENSG00000243477:ENSG00000114378:ENSG00000068001:ENSG00000114735:ENSG00000114738:ENSG00000196455:ENSG00000034533:ENSG00000198585:ENSG00000138246:ENSG00000113971 |
| DEG.twoside | Heart_Atrial_Appendage | 9152 | 44 | 6.85E-05 | 3.70E-03 | ENSG00000115365:ENSG00000021826:ENSG00000100075:ENSG00000100084:ENSG00000070010:ENSG00000215012:ENSG00000093010:ENSG00000099889:ENSG00000128191:ENSG00000099899:ENSG00000099901:ENSG00000099904:ENSG00000099917:ENSG00000114302:ENSG00000178467:ENSG00000178149:ENSG00000198218:ENSG00000177352:ENSG00000188315:ENSG00000114316:ENSG00000145022:ENSG00000145029:ENSG00000173531:ENSG00000164068:ENSG00000173540:ENSG00000176095:ENSG00000185614:ENSG00000182179:ENSG00000164077:ENSG00000001617:ENSG00000186792:ENSG00000243477:ENSG00000007402:ENSG00000114735:ENSG00000114738:ENSG00000088538:ENSG00000196455:ENSG00000017260:ENSG00000034533:ENSG00000114670:ENSG00000198585:ENSG00000138246:ENSG00000113971:ENSG00000004864 |
| DEG.down | Brain_Frontal_Cortex_BA9 | 5015 | 29 | 0.00011 | 5.96E-03 | ENSG00000021826:ENSG00000070371:ENSG00000070010:ENSG00000128191:ENSG00000114302:ENSG00000178537:ENSG00000177479:ENSG00000198218:ENSG00000172037:ENSG00000177352:ENSG00000185909:ENSG00000145020:ENSG00000173402:ENSG00000173531:ENSG00000173540:ENSG00000185614:ENSG00000182179:ENSG00000183763:ENSG00000001617:ENSG00000214706:ENSG00000114378:ENSG00000068001:ENSG00000114735:ENSG00000114738:ENSG00000138246:ENSG00000240303:ENSG00000113971:ENSG00000124664:ENSG00000004864 |
| DEG.down | Brain_Cerebellar_Hemisphere | 2607 | 19 | 0.00014 | 7.34E-03 | ENSG00000183597:ENSG00000178537:ENSG00000172037:ENSG00000177352:ENSG00000185909:ENSG00000173402:ENSG00000173540:ENSG00000182179:ENSG00000164078:ENSG00000001617:ENSG00000214706:ENSG00000114378:ENSG00000068001:ENSG00000114735:ENSG00000114738:ENSG00000114670:ENSG00000113971:ENSG00000124664:ENSG00000004864 |
| DEG.twoside | Brain_Anterior_cingulate_cortex_BA24 | 9021 | 42 | 0.00026 | 1.39E-02 | ENSG00000021826:ENSG00000070371:ENSG00000070010:ENSG00000128191:ENSG00000099899:ENSG00000234409:ENSG00000099917:ENSG00000008300:ENSG00000114302:ENSG00000178537:ENSG00000177479:ENSG00000178149:ENSG00000198218:ENSG00000172037:ENSG00000177352:ENSG00000185909:ENSG00000188315:ENSG00000114316:ENSG00000145020:ENSG00000173402:ENSG00000164061:ENSG00000173531:ENSG00000164068:ENSG00000173540:ENSG00000185614:ENSG00000182179:ENSG00000183763:ENSG00000164076:ENSG00000001617:ENSG00000214706:ENSG00000114378:ENSG00000068001:ENSG00000114735:ENSG00000114738:ENSG00000088538:ENSG00000196455:ENSG00000034533:ENSG00000138246:ENSG00000240303:ENSG00000113971:ENSG00000124664:ENSG00000004864 |
| DEG.down | Brain_Hypothalamus | 5904 | 31 | 0.00035 | 1.89E-02 | ENSG00000070371:ENSG00000215012:ENSG00000128191:ENSG00000099899:ENSG00000099917:ENSG00000114302:ENSG00000178537:ENSG00000177479:ENSG00000178149:ENSG00000198218:ENSG00000172037:ENSG00000177352:ENSG00000114316:ENSG00000145020:ENSG00000173402:ENSG00000173531:ENSG00000164068:ENSG00000173540:ENSG00000185614:ENSG00000182179:ENSG00000001617:ENSG00000214706:ENSG00000114378:ENSG00000068001:ENSG00000114735:ENSG00000114738:ENSG00000196455:ENSG00000138246:ENSG00000240303:ENSG00000113971:ENSG00000004864 |
| DEG.down | Colon_Transverse | 1583 | 13 | 0.0006 | 3.24E-02 | ENSG00000021826:ENSG00000070371:ENSG00000184058:ENSG00000234409:ENSG00000008300:ENSG00000173531:ENSG00000185614:ENSG00000007402:ENSG00000114738:ENSG00000088538:ENSG00000114670:ENSG00000129048:ENSG00000125388 |
| DEG.twoside | Brain_Frontal_Cortex_BA9 | 8147 | 37 | 0.00133 | 7.18E-02 | ENSG00000021826:ENSG00000070371:ENSG00000070010:ENSG00000128191:ENSG00000008300:ENSG00000213672:ENSG00000114302:ENSG00000178537:ENSG00000177479:ENSG00000178467:ENSG00000198218:ENSG00000172037:ENSG00000177352:ENSG00000185909:ENSG00000145020:ENSG00000173402:ENSG00000164061:ENSG00000173531:ENSG00000173540:ENSG00000185614:ENSG00000182179:ENSG00000183763:ENSG00000164076:ENSG00000164077:ENSG00000001617:ENSG00000214706:ENSG00000186792:ENSG00000114378:ENSG00000068001:ENSG00000114735:ENSG00000114738:ENSG00000088538:ENSG00000138246:ENSG00000240303:ENSG00000113971:ENSG00000124664:ENSG00000004864 |
| DEG.down | Brain_Cortex | 5556 | 28 | 0.00148 | 8.01E-02 | ENSG00000021826:ENSG00000070371:ENSG00000070010:ENSG00000128191:ENSG00000114302:ENSG00000178537:ENSG00000177479:ENSG00000198218:ENSG00000172037:ENSG00000177352:ENSG00000185909:ENSG00000145020:ENSG00000173402:ENSG00000173531:ENSG00000173540:ENSG00000185614:ENSG00000182179:ENSG00000001617:ENSG00000214706:ENSG00000114378:ENSG00000068001:ENSG00000114735:ENSG00000114738:ENSG00000138246:ENSG00000240303:ENSG00000113971:ENSG00000124664:ENSG00000004864 |
| DEG.twoside | Liver | 9510 | 41 | 0.00177 | 9.57E-02 | ENSG00000115365:ENSG00000021826:ENSG00000070371:ENSG00000070010:ENSG00000215012:ENSG00000099889:ENSG00000183597:ENSG00000128191:ENSG00000099904:ENSG00000099917:ENSG00000213672:ENSG00000114302:ENSG00000178537:ENSG00000177479:ENSG00000178467:ENSG00000178149:ENSG00000198218:ENSG00000177352:ENSG00000185909:ENSG00000188315:ENSG00000114316:ENSG00000145029:ENSG00000173402:ENSG00000173531:ENSG00000176095:ENSG00000182179:ENSG00000003756:ENSG00000001617:ENSG00000243477:ENSG00000114378:ENSG00000068001:ENSG00000114383:ENSG00000114738:ENSG00000196455:ENSG00000017260:ENSG00000034533:ENSG00000138246:ENSG00000240303:ENSG00000113971:ENSG00000004864:ENSG00000092964 |
| DEG.twoside | Brain_Cerebellum | 8903 | 39 | 0.00185 | 9.97E-02 | ENSG00000115365:ENSG00000100084:ENSG00000215012:ENSG00000099889:ENSG00000183597:ENSG00000128191:ENSG00000099899:ENSG00000099904:ENSG00000234409:ENSG00000008300:ENSG00000213672:ENSG00000114302:ENSG00000178537:ENSG00000178467:ENSG00000172037:ENSG00000177352:ENSG00000185909:ENSG00000188315:ENSG00000145020:ENSG00000145029:ENSG00000164061:ENSG00000164068:ENSG00000173540:ENSG00000176095:ENSG00000182179:ENSG00000164078:ENSG00000001617:ENSG00000186792:ENSG00000243477:ENSG00000114378:ENSG00000068001:ENSG00000007402:ENSG00000114735:ENSG00000114738:ENSG00000088538:ENSG00000113971:ENSG00000125388:ENSG00000124664:ENSG00000004864 |
| DEG.down | Brain_Cerebellum | 2739 | 17 | 0.00196 | 1.06E-01 | ENSG00000183597:ENSG00000114302:ENSG00000178537:ENSG00000172037:ENSG00000177352:ENSG00000185909:ENSG00000173540:ENSG00000182179:ENSG00000164078:ENSG00000001617:ENSG00000114378:ENSG00000068001:ENSG00000114735:ENSG00000114738:ENSG00000113971:ENSG00000124664:ENSG00000004864 |
| DEG.twoside | Brain_Hypothalamus | 8435 | 37 | 0.00259 | 1.40E-01 | ENSG00000070371:ENSG00000215012:ENSG00000128191:ENSG00000099899:ENSG00000099917:ENSG00000008300:ENSG00000114302:ENSG00000178537:ENSG00000177479:ENSG00000178149:ENSG00000198218:ENSG00000172037:ENSG00000177352:ENSG00000114316:ENSG00000145020:ENSG00000173402:ENSG00000164061:ENSG00000173531:ENSG00000164068:ENSG00000173540:ENSG00000185614:ENSG00000182179:ENSG00000164076:ENSG00000001617:ENSG00000214706:ENSG00000114378:ENSG00000068001:ENSG00000007402:ENSG00000114735:ENSG00000114738:ENSG00000088538:ENSG00000196455:ENSG00000114670:ENSG00000138246:ENSG00000240303:ENSG00000113971:ENSG00000004864 |
| DEG.down | Cells_EBV-transformed_lymphocytes | 2383 | 15 | 0.00325 | 1.76E-01 | ENSG00000021826:ENSG00000099904:ENSG00000178467:ENSG00000185909:ENSG00000188315:ENSG00000145020:ENSG00000173402:ENSG00000164068:ENSG00000185614:ENSG00000164078:ENSG00000068001:ENSG00000198585:ENSG00000113971:ENSG00000125388:ENSG00000092964 |
| DEG.twoside | Brain_Cerebellar_Hemisphere | 8908 | 38 | 0.00368 | 1.98E-01 | ENSG00000115365:ENSG00000100084:ENSG00000215012:ENSG00000099889:ENSG00000183597:ENSG00000128191:ENSG00000099904:ENSG00000008300:ENSG00000213672:ENSG00000178537:ENSG00000178467:ENSG00000172037:ENSG00000177352:ENSG00000185909:ENSG00000188315:ENSG00000145020:ENSG00000145029:ENSG00000173402:ENSG00000164061:ENSG00000164068:ENSG00000173540:ENSG00000176095:ENSG00000182179:ENSG00000164078:ENSG00000001617:ENSG00000214706:ENSG00000186792:ENSG00000114378:ENSG00000068001:ENSG00000007402:ENSG00000114735:ENSG00000114738:ENSG00000088538:ENSG00000114670:ENSG00000113971:ENSG00000125388:ENSG00000124664:ENSG00000004864 |
| DEG.down | Liver | 7985 | 35 | 0.00371 | 2.00E-01 | ENSG00000115365:ENSG00000070371:ENSG00000070010:ENSG00000215012:ENSG00000099889:ENSG00000183597:ENSG00000128191:ENSG00000099904:ENSG00000099917:ENSG00000213672:ENSG00000114302:ENSG00000177479:ENSG00000178467:ENSG00000178149:ENSG00000198218:ENSG00000177352:ENSG00000185909:ENSG00000188315:ENSG00000114316:ENSG00000145029:ENSG00000173402:ENSG00000176095:ENSG00000182179:ENSG00000003756:ENSG00000001617:ENSG00000243477:ENSG00000068001:ENSG00000114383:ENSG00000114738:ENSG00000196455:ENSG00000017260:ENSG00000034533:ENSG00000138246:ENSG00000113971:ENSG00000092964 |
| DEG.twoside | Brain_Cortex | 8348 | 36 | 0.00418 | 2.26E-01 | ENSG00000021826:ENSG00000070371:ENSG00000070010:ENSG00000128191:ENSG00000008300:ENSG00000213672:ENSG00000114302:ENSG00000178537:ENSG00000177479:ENSG00000178467:ENSG00000198218:ENSG00000172037:ENSG00000177352:ENSG00000185909:ENSG00000145020:ENSG00000173402:ENSG00000164061:ENSG00000173531:ENSG00000173540:ENSG00000185614:ENSG00000182179:ENSG00000164076:ENSG00000164077:ENSG00000001617:ENSG00000214706:ENSG00000186792:ENSG00000114378:ENSG00000068001:ENSG00000114735:ENSG00000114738:ENSG00000088538:ENSG00000138246:ENSG00000240303:ENSG00000113971:ENSG00000124664:ENSG00000004864 |
| DEG.up | Thyroid | 5687 | 26 | 0.00883 | 4.77E-01 | ENSG00000100084:ENSG00000184058:ENSG00000215012:ENSG00000099889:ENSG00000183597:ENSG00000128191:ENSG00000099899:ENSG00000099904:ENSG00000178467:ENSG00000178149:ENSG00000177352:ENSG00000145022:ENSG00000145020:ENSG00000145029:ENSG00000173531:ENSG00000173540:ENSG00000182179:ENSG00000001617:ENSG00000068001:ENSG00000114735:ENSG00000114738:ENSG00000088538:ENSG00000196455:ENSG00000034533:ENSG00000114670:ENSG00000113971 |
| DEG.down | Ovary | 1521 | 10 | 0.01221 | 6.59E-01 | ENSG00000183597:ENSG00000234409:ENSG00000008300:ENSG00000178537:ENSG00000185614:ENSG00000164078:ENSG00000001617:ENSG00000114378:ENSG00000088538:ENSG00000129048 |
| DEG.down | Kidney_Cortex | 5544 | 25 | 0.01233 | 6.66E-01 | ENSG00000115365:ENSG00000021826:ENSG00000070371:ENSG00000070010:ENSG00000215012:ENSG00000128191:ENSG00000114302:ENSG00000177479:ENSG00000198218:ENSG00000177352:ENSG00000185909:ENSG00000188315:ENSG00000114316:ENSG00000176095:ENSG00000185614:ENSG00000183763:ENSG00000186792:ENSG00000243477:ENSG00000196455:ENSG00000017260:ENSG00000034533:ENSG00000138246:ENSG00000113971:ENSG00000125388:ENSG00000092964 |
| DEG.down | Stomach | 2607 | 14 | 0.01698 | 9.17E-01 | ENSG00000115365:ENSG00000070371:ENSG00000184058:ENSG00000234409:ENSG00000008300:ENSG00000213672:ENSG00000188315:ENSG00000114316:ENSG00000173531:ENSG00000185614:ENSG00000088538:ENSG00000114670:ENSG00000125388:ENSG00000092964 |
| DEG.twoside | Thyroid | 6725 | 28 | 0.02184 | 1.00E+00 | ENSG00000100084:ENSG00000184058:ENSG00000215012:ENSG00000099889:ENSG00000183597:ENSG00000128191:ENSG00000099899:ENSG00000099904:ENSG00000008300:ENSG00000178467:ENSG00000178149:ENSG00000177352:ENSG00000145022:ENSG00000145020:ENSG00000145029:ENSG00000173531:ENSG00000173540:ENSG00000185614:ENSG00000182179:ENSG00000001617:ENSG00000068001:ENSG00000114735:ENSG00000114738:ENSG00000088538:ENSG00000196455:ENSG00000034533:ENSG00000114670:ENSG00000113971 |
| DEG.twoside | Nerve_Tibial | 6774 | 28 | 0.0239 | 1.00E+00 | ENSG00000070371:ENSG00000099889:ENSG00000099899:ENSG00000099904:ENSG00000234409:ENSG00000213672:ENSG00000178467:ENSG00000178149:ENSG00000177352:ENSG00000145020:ENSG00000145029:ENSG00000173531:ENSG00000182179:ENSG00000164078:ENSG00000186792:ENSG00000243477:ENSG00000068001:ENSG00000007402:ENSG00000114738:ENSG00000088538:ENSG00000196455:ENSG00000017260:ENSG00000034533:ENSG00000198585:ENSG00000138246:ENSG00000240303:ENSG00000113971:ENSG00000125388 |
| DEG.down | Esophagus_Mucosa | 3580 | 17 | 0.02606 | 1.00E+00 | ENSG00000070371:ENSG00000099889:ENSG00000183597:ENSG00000213672:ENSG00000178537:ENSG00000178467:ENSG00000145022:ENSG00000145020:ENSG00000145029:ENSG00000173531:ENSG00000114378:ENSG00000088538:ENSG00000114670:ENSG00000198585:ENSG00000113971:ENSG00000125388:ENSG00000092964 |
| DEG.twoside | Artery_Aorta | 4216 | 19 | 0.03038 | 1.00E+00 | ENSG00000070371:ENSG00000184058:ENSG00000099889:ENSG00000183597:ENSG00000234409:ENSG00000213672:ENSG00000145020:ENSG00000145029:ENSG00000173531:ENSG00000185614:ENSG00000182179:ENSG00000001617:ENSG00000114378:ENSG00000114738:ENSG00000088538:ENSG00000138246:ENSG00000240303:ENSG00000113971:ENSG00000129048 |
| DEG.down | Spleen | 2069 | 11 | 0.0356 | 1.00E+00 | ENSG00000021826:ENSG00000070371:ENSG00000184058:ENSG00000114302:ENSG00000173402:ENSG00000164078:ENSG00000007402:ENSG00000088538:ENSG00000114670:ENSG00000129048:ENSG00000125388 |
| DEG.twoside | Colon_Transverse | 3185 | 15 | 0.03859 | 1.00E+00 | ENSG00000021826:ENSG00000070371:ENSG00000184058:ENSG00000234409:ENSG00000008300:ENSG00000173531:ENSG00000185614:ENSG00000164078:ENSG00000007402:ENSG00000114738:ENSG00000088538:ENSG00000114670:ENSG00000129048:ENSG00000125388:ENSG00000124664 |
| DEG.down | Vagina | 647 | 5 | 0.03941 | 1.00E+00 | ENSG00000021826:ENSG00000008300:ENSG00000007402:ENSG00000088538:ENSG00000129048 |
| DEG.up | Nerve_Tibial | 5869 | 24 | 0.04205 | 1.00E+00 | ENSG00000070371:ENSG00000099889:ENSG00000099899:ENSG00000099904:ENSG00000234409:ENSG00000213672:ENSG00000178467:ENSG00000178149:ENSG00000177352:ENSG00000145020:ENSG00000145029:ENSG00000173531:ENSG00000182179:ENSG00000243477:ENSG00000068001:ENSG00000088538:ENSG00000196455:ENSG00000017260:ENSG00000034533:ENSG00000198585:ENSG00000138246:ENSG00000240303:ENSG00000113971:ENSG00000125388 |
| DEG.up | Muscle_Skeletal | 1143 | 7 | 0.0472 | 1.00E+00 | ENSG00000070371:ENSG00000184058:ENSG00000114302:ENSG00000178537:ENSG00000177479:ENSG00000164068:ENSG00000114738 |
| DEG.twoside | Kidney_Cortex | 6871 | 27 | 0.04779 | 1.00E+00 | ENSG00000115365:ENSG00000021826:ENSG00000070371:ENSG00000070010:ENSG00000215012:ENSG00000128191:ENSG00000114302:ENSG00000177479:ENSG00000198218:ENSG00000177352:ENSG00000185909:ENSG00000188315:ENSG00000114316:ENSG00000173531:ENSG00000176095:ENSG00000185614:ENSG00000183763:ENSG00000186792:ENSG00000243477:ENSG00000114378:ENSG00000196455:ENSG00000017260:ENSG00000034533:ENSG00000138246:ENSG00000113971:ENSG00000125388:ENSG00000092964 |
| Enriched gene-sets at nominal p-value <0.05 were reported. | | | | | | |

| **Supplementary Table S8.** Differential gene set enrichment in 30 general tissue types from GTEx. | | | | | | |
| --- | --- | --- | --- | --- | --- | --- |
| Category | GeneSet | N_genes | N_overlap | p | adjP | genes |
| DEG.twoside | Muscle | 7979 | 51 | 3.27E-10 | 9.80E-09 | ENSG00000115365:ENSG00000100075:ENSG00000070371:ENSG00000184058:ENSG00000215012:ENSG00000093010:ENSG00000099889:ENSG00000128191:ENSG00000099899:ENSG00000099901:ENSG00000099904:ENSG00000213672:ENSG00000114302:ENSG00000178537:ENSG00000177479:ENSG00000178467:ENSG00000178149:ENSG00000198218:ENSG00000177352:ENSG00000188315:ENSG00000114316:ENSG00000145022:ENSG00000145020:ENSG00000145029:ENSG00000173531:ENSG00000164068:ENSG00000173540:ENSG00000176095:ENSG00000185614:ENSG00000182179:ENSG00000183763:ENSG00000004534:ENSG00000001617:ENSG00000114353:ENSG00000243477:ENSG00000114378:ENSG00000068001:ENSG00000007402:ENSG00000114735:ENSG00000114738:ENSG00000088538:ENSG00000196455:ENSG00000017260:ENSG00000034533:ENSG00000198585:ENSG00000138246:ENSG00000240303:ENSG00000113971:ENSG00000125388:ENSG00000004864:ENSG00000092964 |
| DEG.down | Muscle | 6836 | 44 | 1.39E-08 | 4.17E-07 | ENSG00000115365:ENSG00000100075:ENSG00000215012:ENSG00000093010:ENSG00000099889:ENSG00000128191:ENSG00000099899:ENSG00000099901:ENSG00000099904:ENSG00000213672:ENSG00000178467:ENSG00000178149:ENSG00000198218:ENSG00000177352:ENSG00000188315:ENSG00000114316:ENSG00000145022:ENSG00000145020:ENSG00000145029:ENSG00000173531:ENSG00000173540:ENSG00000176095:ENSG00000185614:ENSG00000182179:ENSG00000183763:ENSG00000004534:ENSG00000001617:ENSG00000114353:ENSG00000243477:ENSG00000114378:ENSG00000068001:ENSG00000007402:ENSG00000114735:ENSG00000088538:ENSG00000196455:ENSG00000017260:ENSG00000034533:ENSG00000198585:ENSG00000138246:ENSG00000240303:ENSG00000113971:ENSG00000125388:ENSG00000004864:ENSG00000092964 |
| DEG.down | Pancreas | 9586 | 49 | 2.28E-06 | 6.84E-05 | ENSG00000115365:ENSG00000070371:ENSG00000100084:ENSG00000070010:ENSG00000215012:ENSG00000093010:ENSG00000099889:ENSG00000183597:ENSG00000128191:ENSG00000099899:ENSG00000099901:ENSG00000099904:ENSG00000099917:ENSG00000213672:ENSG00000114302:ENSG00000178537:ENSG00000177479:ENSG00000178057:ENSG00000198218:ENSG00000177352:ENSG00000185909:ENSG00000188315:ENSG00000114316:ENSG00000145022:ENSG00000145029:ENSG00000173531:ENSG00000164068:ENSG00000176095:ENSG00000182179:ENSG00000164077:ENSG00000001617:ENSG00000114353:ENSG00000186792:ENSG00000243477:ENSG00000114378:ENSG00000068001:ENSG00000114383:ENSG00000007402:ENSG00000196455:ENSG00000017260:ENSG00000034533:ENSG00000114670:ENSG00000198585:ENSG00000138246:ENSG00000240303:ENSG00000113971:ENSG00000125388:ENSG00000004864:ENSG00000092964 |
| DEG.down | Blood | 6184 | 37 | 2.86E-06 | 8.58E-05 | ENSG00000115365:ENSG00000100075:ENSG00000070371:ENSG00000215012:ENSG00000093010:ENSG00000099889:ENSG00000128191:ENSG00000099901:ENSG00000099904:ENSG00000213672:ENSG00000114302:ENSG00000177479:ENSG00000178467:ENSG00000178252:ENSG00000178149:ENSG00000198218:ENSG00000172037:ENSG00000185909:ENSG00000145020:ENSG00000145029:ENSG00000173402:ENSG00000173531:ENSG00000185614:ENSG00000164077:ENSG00000214706:ENSG00000243477:ENSG00000068001:ENSG00000007402:ENSG00000114735:ENSG00000196455:ENSG00000017260:ENSG00000114686:ENSG00000138246:ENSG00000240303:ENSG00000113971:ENSG00000004864:ENSG00000092964 |
| DEG.twoside | Pancreas | 10331 | 49 | 2.30E-05 | 0.00069032 | ENSG00000115365:ENSG00000070371:ENSG00000100084:ENSG00000070010:ENSG00000215012:ENSG00000093010:ENSG00000099889:ENSG00000183597:ENSG00000128191:ENSG00000099899:ENSG00000099901:ENSG00000099904:ENSG00000099917:ENSG00000213672:ENSG00000114302:ENSG00000178537:ENSG00000177479:ENSG00000178057:ENSG00000198218:ENSG00000177352:ENSG00000185909:ENSG00000188315:ENSG00000114316:ENSG00000145022:ENSG00000145029:ENSG00000173531:ENSG00000164068:ENSG00000176095:ENSG00000182179:ENSG00000164077:ENSG00000001617:ENSG00000114353:ENSG00000186792:ENSG00000243477:ENSG00000114378:ENSG00000068001:ENSG00000114383:ENSG00000007402:ENSG00000196455:ENSG00000017260:ENSG00000034533:ENSG00000114670:ENSG00000198585:ENSG00000138246:ENSG00000240303:ENSG00000113971:ENSG00000125388:ENSG00000004864:ENSG00000092964 |
| DEG.twoside | Heart | 9960 | 46 | 0.00011453 | 0.00343591 | ENSG00000115365:ENSG00000100075:ENSG00000100084:ENSG00000070010:ENSG00000215012:ENSG00000093010:ENSG00000099889:ENSG00000128191:ENSG00000099899:ENSG00000099901:ENSG00000099904:ENSG00000099917:ENSG00000114302:ENSG00000178467:ENSG00000178149:ENSG00000198218:ENSG00000177352:ENSG00000188315:ENSG00000114316:ENSG00000145022:ENSG00000145029:ENSG00000173531:ENSG00000164068:ENSG00000173540:ENSG00000176095:ENSG00000185614:ENSG00000182179:ENSG00000164077:ENSG00000001617:ENSG00000186792:ENSG00000243477:ENSG00000114378:ENSG00000114383:ENSG00000114735:ENSG00000114738:ENSG00000088538:ENSG00000196455:ENSG00000017260:ENSG00000034533:ENSG00000114670:ENSG00000198585:ENSG00000138246:ENSG00000240303:ENSG00000113971:ENSG00000125388:ENSG00000004864 |
| DEG.down | Heart | 9347 | 44 | 0.000118 | 0.0035401 | ENSG00000115365:ENSG00000100075:ENSG00000100084:ENSG00000070010:ENSG00000215012:ENSG00000093010:ENSG00000099889:ENSG00000128191:ENSG00000099899:ENSG00000099901:ENSG00000099904:ENSG00000099917:ENSG00000114302:ENSG00000178467:ENSG00000178149:ENSG00000198218:ENSG00000177352:ENSG00000188315:ENSG00000114316:ENSG00000145022:ENSG00000145029:ENSG00000173531:ENSG00000164068:ENSG00000173540:ENSG00000176095:ENSG00000185614:ENSG00000182179:ENSG00000164077:ENSG00000001617:ENSG00000186792:ENSG00000243477:ENSG00000114383:ENSG00000114735:ENSG00000088538:ENSG00000196455:ENSG00000017260:ENSG00000034533:ENSG00000114670:ENSG00000198585:ENSG00000138246:ENSG00000240303:ENSG00000113971:ENSG00000125388:ENSG00000004864 |
| DEG.twoside | Blood | 8434 | 40 | 0.00027332 | 0.00819957 | ENSG00000115365:ENSG00000100075:ENSG00000070371:ENSG00000093009:ENSG00000215012:ENSG00000093010:ENSG00000099889:ENSG00000183597:ENSG00000128191:ENSG00000099901:ENSG00000099904:ENSG00000213672:ENSG00000114302:ENSG00000177479:ENSG00000178467:ENSG00000178252:ENSG00000178149:ENSG00000198218:ENSG00000172037:ENSG00000185909:ENSG00000188315:ENSG00000145020:ENSG00000145029:ENSG00000173402:ENSG00000173531:ENSG00000185614:ENSG00000164077:ENSG00000214706:ENSG00000243477:ENSG00000068001:ENSG00000007402:ENSG00000114735:ENSG00000196455:ENSG00000017260:ENSG00000114686:ENSG00000138246:ENSG00000240303:ENSG00000113971:ENSG00000004864:ENSG00000092964 |
| DEG.down | Brain | 5503 | 29 | 0.00056083 | 0.01682482 | ENSG00000070371:ENSG00000070010:ENSG00000183597:ENSG00000099917:ENSG00000114302:ENSG00000178537:ENSG00000177479:ENSG00000178149:ENSG00000198218:ENSG00000172037:ENSG00000177352:ENSG00000185909:ENSG00000173402:ENSG00000164062:ENSG00000173531:ENSG00000173540:ENSG00000185614:ENSG00000182179:ENSG00000001617:ENSG00000214706:ENSG00000114378:ENSG00000068001:ENSG00000114735:ENSG00000114738:ENSG00000196455:ENSG00000138246:ENSG00000240303:ENSG00000113971:ENSG00000004864 |
| DEG.twoside | Liver | 9510 | 41 | 0.00177281 | 0.05318443 | ENSG00000115365:ENSG00000021826:ENSG00000070371:ENSG00000070010:ENSG00000215012:ENSG00000099889:ENSG00000183597:ENSG00000128191:ENSG00000099904:ENSG00000099917:ENSG00000213672:ENSG00000114302:ENSG00000178537:ENSG00000177479:ENSG00000178467:ENSG00000178149:ENSG00000198218:ENSG00000177352:ENSG00000185909:ENSG00000188315:ENSG00000114316:ENSG00000145029:ENSG00000173402:ENSG00000173531:ENSG00000176095:ENSG00000182179:ENSG00000003756:ENSG00000001617:ENSG00000243477:ENSG00000114378:ENSG00000068001:ENSG00000114383:ENSG00000114738:ENSG00000196455:ENSG00000017260:ENSG00000034533:ENSG00000138246:ENSG00000240303:ENSG00000113971:ENSG00000004864:ENSG00000092964 |
| DEG.twoside | Brain | 8711 | 38 | 0.00240413 | 0.07212392 | ENSG00000115365:ENSG00000070371:ENSG00000070010:ENSG00000183597:ENSG00000099917:ENSG00000008300:ENSG00000114302:ENSG00000178537:ENSG00000177479:ENSG00000178467:ENSG00000178149:ENSG00000198218:ENSG00000172037:ENSG00000177352:ENSG00000185909:ENSG00000173402:ENSG00000164061:ENSG00000164062:ENSG00000173531:ENSG00000173540:ENSG00000185614:ENSG00000182179:ENSG00000164076:ENSG00000001617:ENSG00000214706:ENSG00000186792:ENSG00000114378:ENSG00000068001:ENSG00000007402:ENSG00000114735:ENSG00000114738:ENSG00000088538:ENSG00000196455:ENSG00000138246:ENSG00000240303:ENSG00000113971:ENSG00000004864:ENSG00000092964 |
| DEG.down | Liver | 7985 | 35 | 0.00370878 | 0.11126355 | ENSG00000115365:ENSG00000070371:ENSG00000070010:ENSG00000215012:ENSG00000099889:ENSG00000183597:ENSG00000128191:ENSG00000099904:ENSG00000099917:ENSG00000213672:ENSG00000114302:ENSG00000177479:ENSG00000178467:ENSG00000178149:ENSG00000198218:ENSG00000177352:ENSG00000185909:ENSG00000188315:ENSG00000114316:ENSG00000145029:ENSG00000173402:ENSG00000176095:ENSG00000182179:ENSG00000003756:ENSG00000001617:ENSG00000243477:ENSG00000068001:ENSG00000114383:ENSG00000114738:ENSG00000196455:ENSG00000017260:ENSG00000034533:ENSG00000138246:ENSG00000113971:ENSG00000092964 |
| DEG.down | Colon | 1378 | 10 | 0.00633741 | 0.19012222 | ENSG00000021826:ENSG00000070371:ENSG00000184058:ENSG00000234409:ENSG00000173531:ENSG00000185614:ENSG00000114378:ENSG00000007402:ENSG00000114738:ENSG00000129048 |
| DEG.up | Thyroid | 5687 | 26 | 0.00882731 | 0.2648194 | ENSG00000100084:ENSG00000184058:ENSG00000215012:ENSG00000099889:ENSG00000183597:ENSG00000128191:ENSG00000099899:ENSG00000099904:ENSG00000178467:ENSG00000178149:ENSG00000177352:ENSG00000145022:ENSG00000145020:ENSG00000145029:ENSG00000173531:ENSG00000173540:ENSG00000182179:ENSG00000001617:ENSG00000068001:ENSG00000114735:ENSG00000114738:ENSG00000088538:ENSG00000196455:ENSG00000034533:ENSG00000114670:ENSG00000113971 |
| DEG.down | Ovary | 1521 | 10 | 0.01220929 | 0.36627884 | ENSG00000183597:ENSG00000234409:ENSG00000008300:ENSG00000178537:ENSG00000185614:ENSG00000164078:ENSG00000001617:ENSG00000114378:ENSG00000088538:ENSG00000129048 |
| DEG.up | Skin | 3125 | 16 | 0.01636585 | 0.49097563 | ENSG00000021826:ENSG00000093009:ENSG00000184058:ENSG00000185838:ENSG00000215012:ENSG00000178149:ENSG00000177352:ENSG00000185614:ENSG00000183763:ENSG00000164078:ENSG00000001617:ENSG00000114378:ENSG00000114738:ENSG00000017260:ENSG00000129048:ENSG00000124664 |
| DEG.down | Stomach | 2607 | 14 | 0.01698304 | 0.50949123 | ENSG00000115365:ENSG00000070371:ENSG00000184058:ENSG00000234409:ENSG00000008300:ENSG00000213672:ENSG00000188315:ENSG00000114316:ENSG00000173531:ENSG00000185614:ENSG00000088538:ENSG00000114670:ENSG00000125388:ENSG00000092964 |
| DEG.twoside | Thyroid | 6725 | 28 | 0.02184192 | 0.6552575 | ENSG00000100084:ENSG00000184058:ENSG00000215012:ENSG00000099889:ENSG00000183597:ENSG00000128191:ENSG00000099899:ENSG00000099904:ENSG00000008300:ENSG00000178467:ENSG00000178149:ENSG00000177352:ENSG00000145022:ENSG00000145020:ENSG00000145029:ENSG00000173531:ENSG00000173540:ENSG00000185614:ENSG00000182179:ENSG00000001617:ENSG00000068001:ENSG00000114735:ENSG00000114738:ENSG00000088538:ENSG00000196455:ENSG00000034533:ENSG00000114670:ENSG00000113971 |
| DEG.twoside | Nerve | 6774 | 28 | 0.02390169 | 0.71705068 | ENSG00000070371:ENSG00000099889:ENSG00000099899:ENSG00000099904:ENSG00000234409:ENSG00000213672:ENSG00000178467:ENSG00000178149:ENSG00000177352:ENSG00000145020:ENSG00000145029:ENSG00000173531:ENSG00000182179:ENSG00000164078:ENSG00000186792:ENSG00000243477:ENSG00000068001:ENSG00000007402:ENSG00000114738:ENSG00000088538:ENSG00000196455:ENSG00000017260:ENSG00000034533:ENSG00000198585:ENSG00000138246:ENSG00000240303:ENSG00000113971:ENSG00000125388 |
| DEG.down | Kidney | 5412 | 23 | 0.03167529 | 0.95025857 | ENSG00000115365:ENSG00000021826:ENSG00000070371:ENSG00000070010:ENSG00000215012:ENSG00000128191:ENSG00000114302:ENSG00000177479:ENSG00000198218:ENSG00000177352:ENSG00000188315:ENSG00000114316:ENSG00000185614:ENSG00000183763:ENSG00000186792:ENSG00000243477:ENSG00000196455:ENSG00000017260:ENSG00000034533:ENSG00000138246:ENSG00000113971:ENSG00000125388:ENSG00000092964 |
| DEG.down | Spleen | 2069 | 11 | 0.03559843 | 1 | ENSG00000021826:ENSG00000070371:ENSG00000184058:ENSG00000114302:ENSG00000173402:ENSG00000164078:ENSG00000007402:ENSG00000088538:ENSG00000114670:ENSG00000129048:ENSG00000125388 |
| DEG.down | Vagina | 647 | 5 | 0.03941349 | 1 | ENSG00000021826:ENSG00000008300:ENSG00000007402:ENSG00000088538:ENSG00000129048 |
| DEG.twoside | Skin | 5256 | 22 | 0.04144981 | 1 | ENSG00000021826:ENSG00000093009:ENSG00000184058:ENSG00000185838:ENSG00000215012:ENSG00000099889:ENSG00000008300:ENSG00000178467:ENSG00000178149:ENSG00000177352:ENSG00000185614:ENSG00000183763:ENSG00000164078:ENSG00000001617:ENSG00000114378:ENSG00000007402:ENSG00000114738:ENSG00000088538:ENSG00000017260:ENSG00000129048:ENSG00000125388:ENSG00000124664 |
| DEG.up | Nerve | 5869 | 24 | 0.04204588 | 1 | ENSG00000070371:ENSG00000099889:ENSG00000099899:ENSG00000099904:ENSG00000234409:ENSG00000213672:ENSG00000178467:ENSG00000178149:ENSG00000177352:ENSG00000145020:ENSG00000145029:ENSG00000173531:ENSG00000182179:ENSG00000243477:ENSG00000068001:ENSG00000088538:ENSG00000196455:ENSG00000017260:ENSG00000034533:ENSG00000198585:ENSG00000138246:ENSG00000240303:ENSG00000113971:ENSG00000125388 |
| DEG.up | Muscle | 1143 | 7 | 0.04719654 | 1 | ENSG00000070371:ENSG00000184058:ENSG00000114302:ENSG00000178537:ENSG00000177479:ENSG00000164068:ENSG00000114738 |
| Enriched gene-sets at nominal p-value <0.05 were reported. | | | | | | |

| **Supplementary Table S9.** Colocalization of *RNF123* locus with muscle skeletal eQTL signals. | | | | | | | | | | | | |
| --- | --- | --- | --- | --- | --- | --- | --- | --- | --- | --- | --- | --- |
| **Locus** | | **CWP GWAS** | | | | **Muscle eQTL** | | | | **LD** | **PP** | |
| GWAS | eQTL | Lead SNP | MAF | N | P | Lead eSNP | MAF | N | P | r^2^ | PP3 | PP4 |
| *RNF123* | *CDHR4* | rs1491985 | 0.18 | 249843 | 1.6E-08 | rs6809879 | 019 | 706 | 3.1E-08 | 1 | 0.07 | 0.93 |
| CWP, chronic widespread pain; DRG, dorsal root ganglion; eQTL, expression quantitative trait loci; MAF, minor allele frequency; N, sample size; P, p-value; LD, linkage disequilibrium; r^2^, the pairwise LD between the lead GWAS SNP, and the lead eSNP; PP, posterior probability; PP3, the posterior probabilities for having separate variants for both traits, PP4, the posterior probabilities for having shared SNP between two traits. eQTL SNP with lowest p-value was reported. | | | | | | | | | | | | |

| **Supplementary Table S10.** Colocalization of *RNF123* locus with DRG eQTL signals at exon-level. | | | | | | | | | | | | |
| --- | --- | --- | --- | --- | --- | --- | --- | --- | --- | --- | --- | --- |
| **Locus** | | **CWP GWAS** | | | | **DRG eQTL** | | | | **LD** | **PP** | |
| GWAS | eQTL | Lead SNP | MAF | N | P | Lead eSNP | MAF | N | P | r^2^ | PP3 | PP4 |
| *RNF123* | *APEH* | rs1491985 | 0.18 | 249843 | 3.40E-08 | rs13093525 | 0.16 | 214 | 1.32E-06 | 1 | 0.01 | 0.72 |
| CWP, chronic widespread pain; DRG, dorsal root ganglion; eQTL, expression quantitative trait loci; MAF, minor allele frequency; N, sample size; P, p-value; LD, linkage disequilibrium; r^2^, the pairwise LD between the lead GWAS SNP, and the lead eSNP; PP, posterior probability; PP3, the posterior probabilities for having separate variants for both traits, PP4, the posterior probabilities for having shared SNP between two traits. eQTL SNP with lowest p-value was reported. | | | | | | | | | | | | |
